# Mapping the Interplay of Atrial Fibrillation, Brain Structure and Cognitive Dysfunction

**DOI:** 10.1101/2023.11.13.23298431

**Authors:** Marvin Petersen, Céleste Chevalier, Felix L. Naegele, Thies Ingwersen, Amir Omidvarnia, Felix Hoffstaedter, Kaustubh Patil, Simon B. Eickhoff, Renate B. Schnabel, Paulus Kirchhof, Eckhard Schlemm, Bastian Cheng, Götz Thomalla, Märit Jensen

## Abstract

Atrial fibrillation (AF) is associated with an elevated risk of cognitive impairment and dementia. The investigation of the cognitive sequelae and alterations of brain structure linked to AF is crucial to help address ensuing health care needs. In this study, we conducted a comprehensive neuropsychological and neuroimaging analysis of 1335 stroke-free individuals with AF (30% women, average age 69.1 years) and compared them with 2683 demographically and cardiovascular risk-matched controls (31% women, average age 69.1 years). Primary study outcomes were neuropsychological test scores and advanced magnetic resonance imaging (MRI) measures of gray matter morphology, gray and white matter microstructure, and white matter hyperintensity (WMH) load. Our analysis identified deficits in attention/executive function, information processing speed and reasoning in individuals with AF. These cognitive impairments were accompanied by a complex imaging profile suggestive of small vessel pathology: (1) reduced cortical thickness and gray matter volume in areas including primary motor, somatosensory and visual cortices as well as the orbitofrontal, lateral prefrontal, posterior insular, and temporal cortices; (2) increased extracellular free-water content in the anterior cingulate, insula, medial prefrontal cortex, medial temporal lobe, and precuneus; (3) widespread microstructural anomalies in the cerebral white matter, marked by lower fractional anisotropy (FA), higher mean diffusivity (MD) and extracellular free-water, and a higher burden of markers of small vessel disease (WMH load and peak width of skeletonized mean diffusivity). Crucially, brain structural differences statistically mediated the link between AF and cognitive performance. By integrating a multimodal analysis approach with extensive clinical and MRI data, our study highlights small vessel pathology as a possible unifying link between AF, cognitive decline and abnormal brain structure. These insights can inform diagnostic approaches and motivate the ongoing implementation of effective therapeutic strategies.

## Introduction

The association between cardiovascular health and cognitive function has gained increasing attention. Atrial fibrillation (AF), the most common cardiac arrhythmia, affects more than 37 million people worldwide, making it a substantial public health concern.^1^ Several meta-analyses indicate that AF is associated with incidence of cognitive decline and dementia even when accounting for prevalent stroke and shared risk factors.^2–5^ Given that treatments can alter the progression of AF, comprehending its impact on the brain is vital for effective prevention and management of cognitive sequelae.

Mechanistic models have been proposed to explain the connection between AF and disorders of cognition. The connection is considered to arise from AF promoting a prothrombotic and pro-inflammatory environment, reduced cardiac output, and subsequent cerebral perturbations like hypoperfusion, inflammation, blood-brain barrier leakage, endothelial dysfunction and small vessel pathology.^6–8^

Magnetic resonance imaging (MRI) allows for mapping structural brain injury which potentially mediates cognitive effects found in AF. Although many studies suggest a link of AF with lower global brain volume and increased small vessel disease burden, the understanding of the association is still limited.^6,9,10^ Existing research often includes AF individuals with concurrent ischemic stroke, potentially confounding results. Studies also largely focus on global brain structural measures, disregarding the potential importance of regionally specific changes. Lastly, small sample sizes in many studies may have led to inconsistent findings.

We argue that for a better understanding of the interplay between AF, cognition, and brain structure, analyses are needed that unify (1) large-scale, population-based cognitive and MRI data to address confounding and ensure reproducibility, (2) broad cognitive phenotyping and (3) advanced neuroimaging techniques to comprehensively characterize the pathomechanistic correlates of AF.^11,12^

Tapping into these research needs, our study aims to advance the understanding of AF-related cognitive impairments by investigating an AF sample from the UK Biobank in a case-control design, leveraging broad cognitive phenotyping and advanced neuroimaging markers of tissue macro- and microstructure on global and regional scales.

## Materials and methods

### Study population

We examined cross-sectional clinical and imaging data from the UK Biobank (age 45-80 years).^13^ To reduce confounding effects, participants with either a history or a current diagnosis of neurological or psychiatric diseases including history of stroke and dementia were excluded (*supplementary table S1*, http://biobank.ndph.ox.ac.uk/showcase/coding.cgi?id=6). Individuals with AF were identified based on ICD-10 diagnoses. A healthy control sample was compiled performing a 1:2 propensity score matching specifically accounting for confounders known to affect cognitive performance as well as anatomical and diffusion MR imaging indices. Matching criteria included age, sex, education, systolic blood pressure, smoking behavior as well as blood cholesterol and glucose levels, using the matchit package in R (v4.3.3).^14^

### Ethics approval

The UKB was ethically approved by the North West Multi-Centre Research Ethics Committee (MREC) and written informed consent was obtained from all participants. Details on the UKB Ethics and Governance framework are provided online (https://www.ukbiobank.ac.uk/media/0xsbmfmw/egf.pdf).^15^ Data usage is covered by a vote of the ethics committee of the medical faculty of the Heinrich Heine University Düsseldorf.

### Cognitive assessments

The UKB uses computerized versions of established tests to evaluate multiple cognitive domains. Here we investigated cognitive scores of attention and executive function (Tower Rearranging Test, Trail Making Tests part B), processing speed (Reaction Time Test, Symbol Digit Substitution Test, Trail Making Tests part A), memory (Numeric Memory Test, Paired Associate Learning Test, Prospective Memory Test), and reasoning (Fluid Intelligence Test, Matrix Pattern Completion Test).^16^ Detailed descriptions of the individual tests can be found elsewhere.^17^ Results on the Trail making Tests and the Reaction Time Test were inverted to ensure that high scores correspond with better cognitive performance across all tests. Within the four domains, respective tests were z-scored and averaged to obtain domain scores as has been done previously.^18^

### Brain imaging

The full UKB neuroimaging protocol can be found online (https://biobank.ctsu.ox.ac.uk/crystal/crystal/docs/brain_mri.pdf).^13^ In brief, 3D T1-weighted rapid acquisition gradient-echo sequence (MPRAGE, 1x1x1mm, 192x256x256, repetition time = 2000 ms, echo time = 2.01 ms) and multi-shell diffusion weighted imaging (2x2x2mm, 104x104x72, repetition time = 3600ms, echo time = 92 ms, 50 diffusion-encoding directions - 50x b=1000 s/mm^2^ and 50x b=2000 s/mm^2^) were acquired on a 3T Siemens Skyra MRI scanner (Siemens, Erlangen, Germany).

A summary of the imaging markers obtained for both gray and white matter is depicted in Figure 1. For an in-depth explanation of image preprocessing, the calculation of macro- and microstructural indices, and quality assurance see *supplementary text S2*.

**Figure 1.**
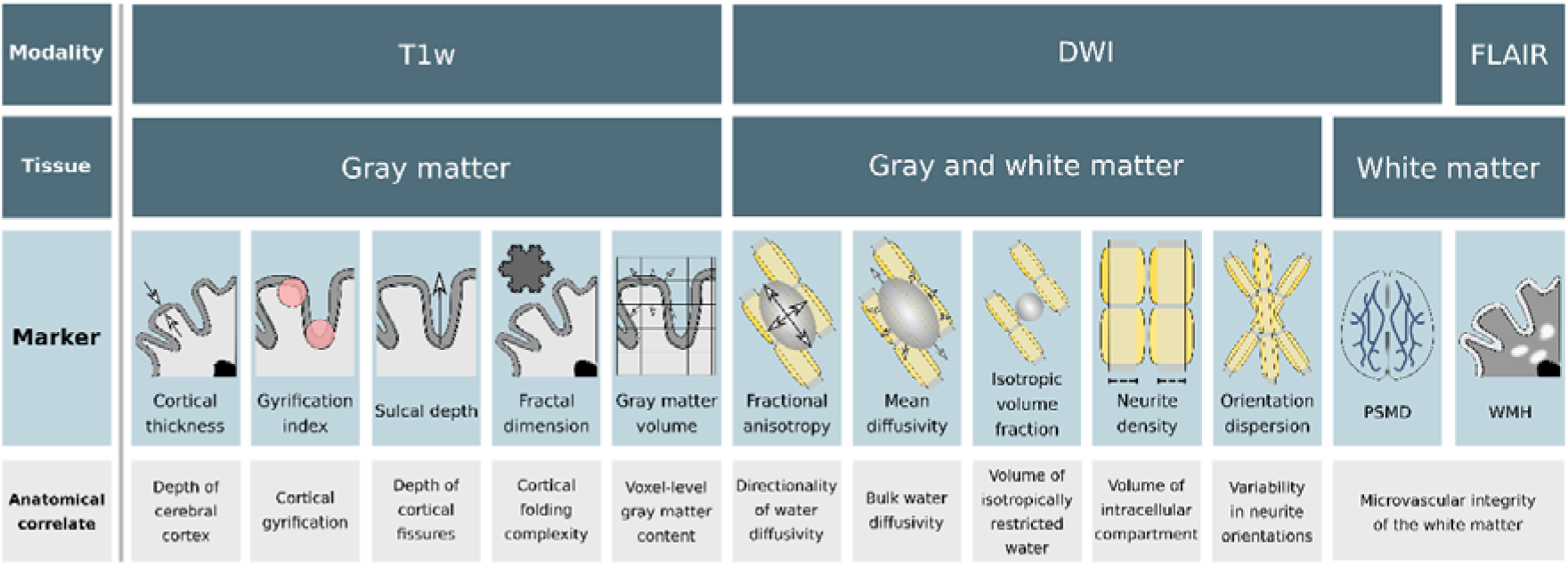
Schematic illustration of the investigated imaging markers. To assess the cerebral gray and white matter, micro- and macrostructural imaging markers were derived. In the first row, the schematic outlines the specific imaging sequences employed to derive the imaging markers. These sequences form the basis for the analysis of the various cerebral structures. The second row delineates the tissues from which the imaging markers were extracted. The third row provides diagrammatic representations of the markers, detailing their specific definitions and functions: cortical thickness is defined as the distance between the pial surface and the boundary between white matter and gray matter; the gyrification index quantifies the folding complexity within the cerebral cortex; sulcal depth measures the depth of the brain’s sulci; fractal dimension informs about the complexity and regularity of cortical folding; gray matter volume estimations are based on voxel-based morphometry utilizing local intensity and registration information to serve as a proxy for brain tissue composition; fractional anisotropy (FA) assesses the directional bias of diffusion; mean diffusivity (MD) denotes the rate of molecular diffusion; the isotropic volume fraction indicates the proportion of isotropic diffusion, reflecting the content of free extracellular water within brain tissue; the neurite density index represents the proportion of water diffusion within neurites, indicative of cellularity and tissue density; orientation dispersion measures the variability in the primary directions of water diffusion, reflecting the coherence or dispersion of neurite orientations. PSMD is a diffusion metric that gauges white matter integrity by mirroring the distribution of mean diffusivity along the white matter skeleton. WMH load refers to the amount of T2-hyperintense lesions, indicative of abnormalities in the white matter resulting from small vessel pathology. Anatomical correlates of the respective imaging markers are described in the fourth row. Modified from *Petersen et al. 2023*.^12^ *Abbreviations*: DWI = diffusion-weighted imaging, FLAIR = fluid-attenuated inversion recovery, PSMD = peak width of skeletonized mean diffusivity, T1w = T1-weighted imaging, WMH = white matter hyperintensity.

Based on T1w images, imaging markers of cortical macrostructure, were computed with the Computational Anatomy Toolbox for SPM (CAT12).^19^ Macrostructural measures inform about large-scale anatomy and geometric characteristics of the brain. Cortical thickness (CT) was measured as the shortest difference between the pial surface and the boundary between white matter and gray matter.^20^ Three measures of cortical folding geometry were obtained: (1) the gyrification index measuring the amount of gyrification, (2) the sulcal depth measuring the depth of the brain’s sulci and (3) the fractal dimension (FD) informing about complexity and regularity of cortical folding.^21–23^ Leveraging voxel-based morphometry (VBM), local intensity and registration information were used to obtain gray matter volume estimates as a proxy of cortical tissue volume.^24,25^ For the statistical analysis, gray matter macrostructural measures were averaged across the whole brain (global level), as well as averaged within Schaefer400 atlas regions of interests (ROI).^26^

Based on DWI, voxel-level imaging markers of tissue microstructure were obtained, i.e., markers representing the underlying organization and arrangement of cells, neurites, and other microscopic components within brain tissue. Following DWI preprocessing, conventional diffusion tensor imaging (DTI) markers of white matter microstructure, i.e., fractional anisotropy (FA) and mean diffusivity (MD), were derived which have been extensively used in neuroscientific and neuropsychological research.^27,28^ In addition, neurite orientation dispersion and density imaging (NODDI) was employed to obtain diffusion measures with higher tissue specificity.^29^ NODDI models (1) the neurite density index (also intracellular volume fraction, ICVF) representing the proportion of water diffusion within neurites, indicative of cellularity and tissue density, (2) isotropic volume fraction (ISOVF) indicating the proportion of isotropic diffusion which quantifies the extracellular free-water content within brain tissue, as well as (3) the orientation dispersion measuring the variability in the primary directions of water diffusion reflecting the coherence or dispersion of neurite orientations. For further statistical analysis, voxel-level microstructural markers were averaged in cortical Schaefer400 atlas regions, as well as 70 predefined anatomical white matter tracts of the normative HCP-842 tractography atlas.^26,30(p201)^ In addition, microstructural markers were projected on a representative skeleton of the entire white matter derived by tract-based spatial statistics (TBSS).^31^ To obtain global markers of gray matter and white matter microstructure, the measures were averaged across the whole gray matter and all voxels of the representative white matter skeleton, respectively.

Finally, the peak width of skeletonized mean diffusivity (PSMD) as well as normalized volumes of white matter hyperintensities (WMH load) were obtained as surrogate markers of microvascular injury in the white matter.^32–34^

### Statistical analysis

All statistical analyses were conducted in python 3.9.1 as well as FSL’s Permutation Analysis of Linear Models (PALM) based on Matlab v.2021b.^35,36^ Statistical tests were two-sided, with a *P*<0.05 as significance threshold. To account for multiple comparisons *P-*values were adjusted via false discovery rate correction.

#### Demographic data

Sample characteristics were compared between AF individuals and healthy controls using *χ2*-tests (binary) and two-sample t-tests (continuous). Clinical variables were compared between groups in separate analyses of covariance (ANCOVA) adjusted for age, sex, education and cardiovascular risk factors.

#### Imaging

Statistical analysis of imaging parameters was performed in two stages. First, global measures, i.e., mean cortical macrostructural markers, mean cortical microstructural markers, mean skeletonized microstructural parameters, WMH load and PSMD, were compared between individuals with AF and healthy controls in separate ANCOVAs, adjusted for age, sex, education and cardiovascular risk. In the case of gray matter volume, total intracranial volume served as an additional covariate.

In a mediation analysis, we tested whether the relationship of AF and cognitive domain scores was mediated by global imaging parameters.^37^ Only scores of cognitive domains that significantly differed between groups were considered. A mediation analysis decomposes the total effect of AF on a cognitive domain performance into two components: (1) the direct, i.e., non-mediated, effect of AF on cognitive domain scores, and (2) the indirect effect, i.e., the portion of the effect that can be attributed to the global imaging parameters. An indirect effect was considered to mediate the relationship between AF and cognition when AF was significantly associated to the mediator, the mediator was significantly associated to the cognitive domain and the link between AF and cognitive domain was reduced (partial mediation) or became non-significant (full mediation) when controlling for the mediator. The presence of a significant mediating effect was determined using bootstrapping (n_bootstrap_=5000). Models were adjusted for age, sex, education and cardiovascular risk.

Next, we examined spatial patterns of brain structural changes linked to AF. We performed region of interest (ROI)-level permutation-based (n_permutation_=5000) testing for two-sided group differences of cortical macrostructural and microstructural markers averaged in Schaefer400 atlas regions as well as white matter microstructural markers in 70 predefined anatomical white matter tracts, respectively. In addition, we performed whole-brain voxel-wise testing of skeletonized microstructural markers conducting TBSS.^31^ TBSS was performed based on the same design matrices as in the ROI-level analysis, i.e. performing non-parametric (n_permutation_=5000) two-sided group-comparisons between cases and controls, with threshold-free cluster enhancement (TFCE) and false discovery-rate correction.^38^

### Sensitivity analysis

We conducted a sensitivity analysis to ascertain the independence of our results from AF-related comorbidities considered to impact cognitive performance and brain health. To address this, we controlled for the diagnosis of comorbidities alongside demographics and cardiovascular risk factors in the statistical models comparing cognitive domain scores and global imaging markers. The comorbidities considered encompassed atherosclerotic heart disease, congestive heart failure, hyperthyroidism, diabetes mellitus type II, alcohol abuse, and chronic obstructive sleep apnea.

### Data availability

UK Biobank data can be obtained via its standardized data access procedure (https://www.ukbiobank.ac.uk/).

## Results

### Sample characteristics

After sample selection, quality assessment and matching, the final analysis sample included 1335 individuals with AF (30% female, mean age 69.1 years) and 2683 matched controls (31% female, mean age 69.1 years; see *table 1*). For a flow chart on the sample selection procedure and balance plots detailing matching results refer to supplementary *figures S3 and S4*. After matching, groups were comparable in terms of age, sex, years of education and cardiovascular risk factors. Information on AF-related comorbidities is displayed in *supplementary table S5*.

**Table 1.** Sample characteristics of AF individuals and matched controls.

| Measure<sup>a</sup> | Atrial fibrillation<sup>a</sup> | Matched controls<sup>a</sup> | $P_{uncorr}$<sup>b</sup> | $P_{FDR}$<sup>c</sup> | Cohen's $d$ or $\chi^2$ |
| --- | --- | --- | --- | --- | --- |
| Age, years | 69.06 ± 6.60 (1335) | 69.05 ± 6.37 (2683) | 0.989 | 0.989 | <0.01 |
| Women, n, % | 404, 30.26 (1335) | 830, 30.94 (2683) | 0.690 | 0.989 | 0.16 |
| Education (ISCED) | 4.29 ± 1.55 (1335) | 4.30 ± 1.49 (2683) | 0.849 | 0.989 | 0.01 |
| Systolic blood pressure, mmHg | 142.06 ± 19.11 (998) | 142.83 ± 18.57 (2161) | 0.288 | 0.721 | 0.04 |
| Diastolic blood pressure, mmHg | 79.57 ± 11.39 (979) | 78.51 ± 10.53 (2104) | 0.014 | 0.072 | -0.10 |
| Current smokers, n, % | 26, 1.95 (1320) | 52, 1.94 (2657) | 0.920 | 0.989 | 0.01 |
| Blood cholesterol, mmol/L | 5.53 ± 1.10 (1257) | 5.54 ± 1.08 (2497) | 0.959 | 0.989 | <0.01 |
| High density lipoprotein, mmol/L | 1.39 ± 0.36 (1142) | 1.41 ± 0.36 (2289) | 0.069 | 0.229 | 0.07 |
| Low density lipoprotein, mmol/L | 3.46 ± 0.84 (1255) | 3.47 ± 0.83 (2490) | 0.964 | 0.989 | <0.01 |
| Blood glucose, mmol/L | 5.08 ± 1.09 (1143) | 5.08 ± 1.06 (2287) | 0.987 | 0.989 | <0.01 |

*Abbreviations:* ISCED – International Standard Classification of Education
<sup>a</sup>Presented as mean ± SD (n of datapoints)
<sup>b</sup>Uncorrected *P*-values of t-tests or $\chi^2$ tests
<sup>c</sup>False discovery rate-corrected *P*-values of t-tests or $\chi^2$ tests

### Cognitive function

Individuals with AF showed worse test performance for the cognitive domains of attention / executive function (mean ± standard deviation [SD], -0.058 ± 0.851 vs. 0.039 ± 0.820, Cohen’s *d* = 0.12, *P_FDR_* = 0.012) and reasoning (mean ± SD, -0.085 ± 0.837 vs. 0.049 ± 0.838, Cohen’s *d* = 0.16, *P_FDR_* <0.001) than matched controls (see *table 2*). Domain scores of memory function and information processing speed showed no significant differences between groups. These results remained stable when additionally controlling for AF comorbidities (*supplementary table S6)*. On the level of individual cognitive tests of information processing speed, the AF group showed lower performance in the Symbol Digit Substitution Test (mean ± SD, 16.68 ± 5.08 vs. 17.28 ± 5.04, Cohen’s *d* = 0.12, *P_FDR_* = 0.034). More details on individual cognitive test performances can be found in *supplementary table S7*.

**Table 2.**
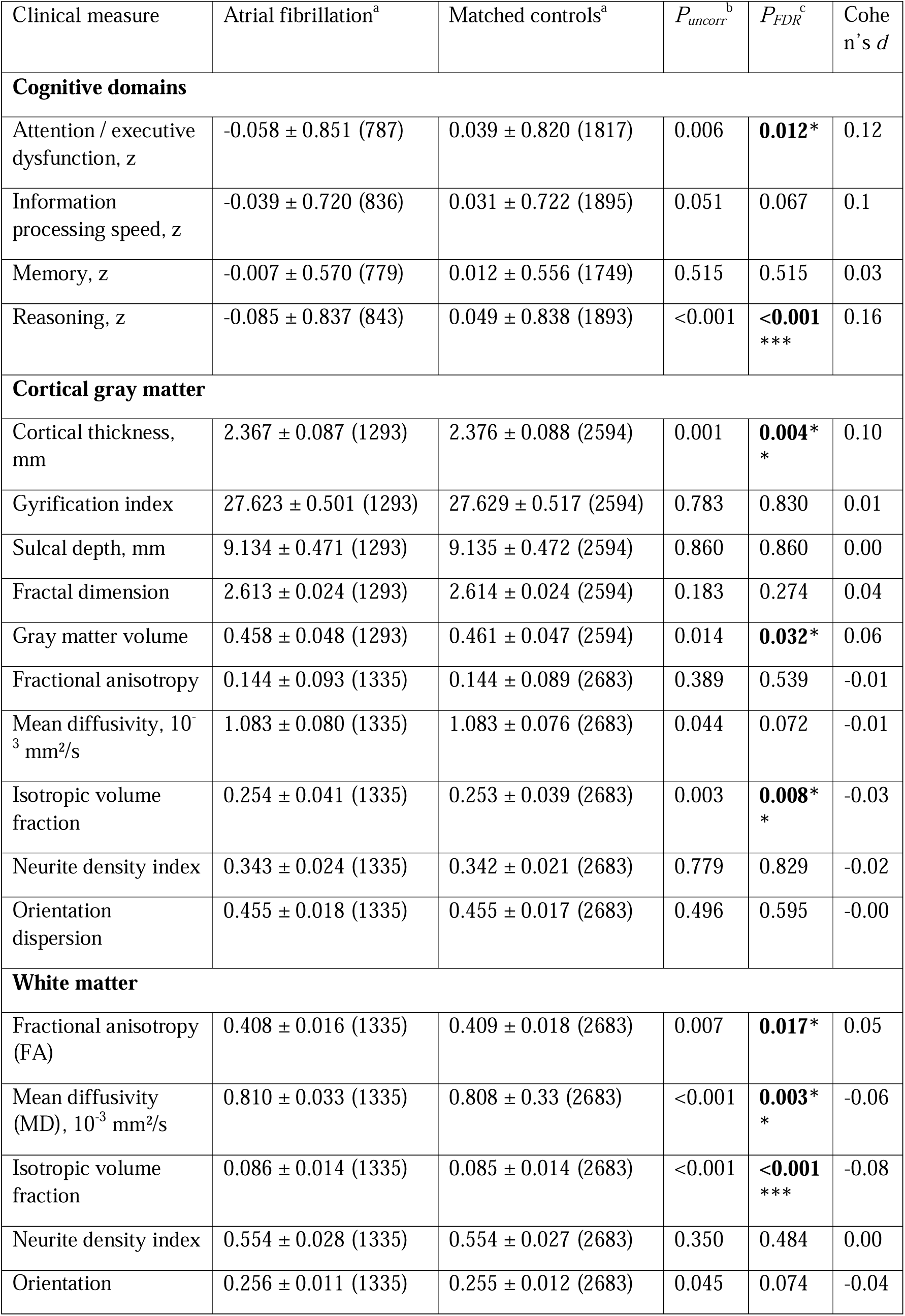
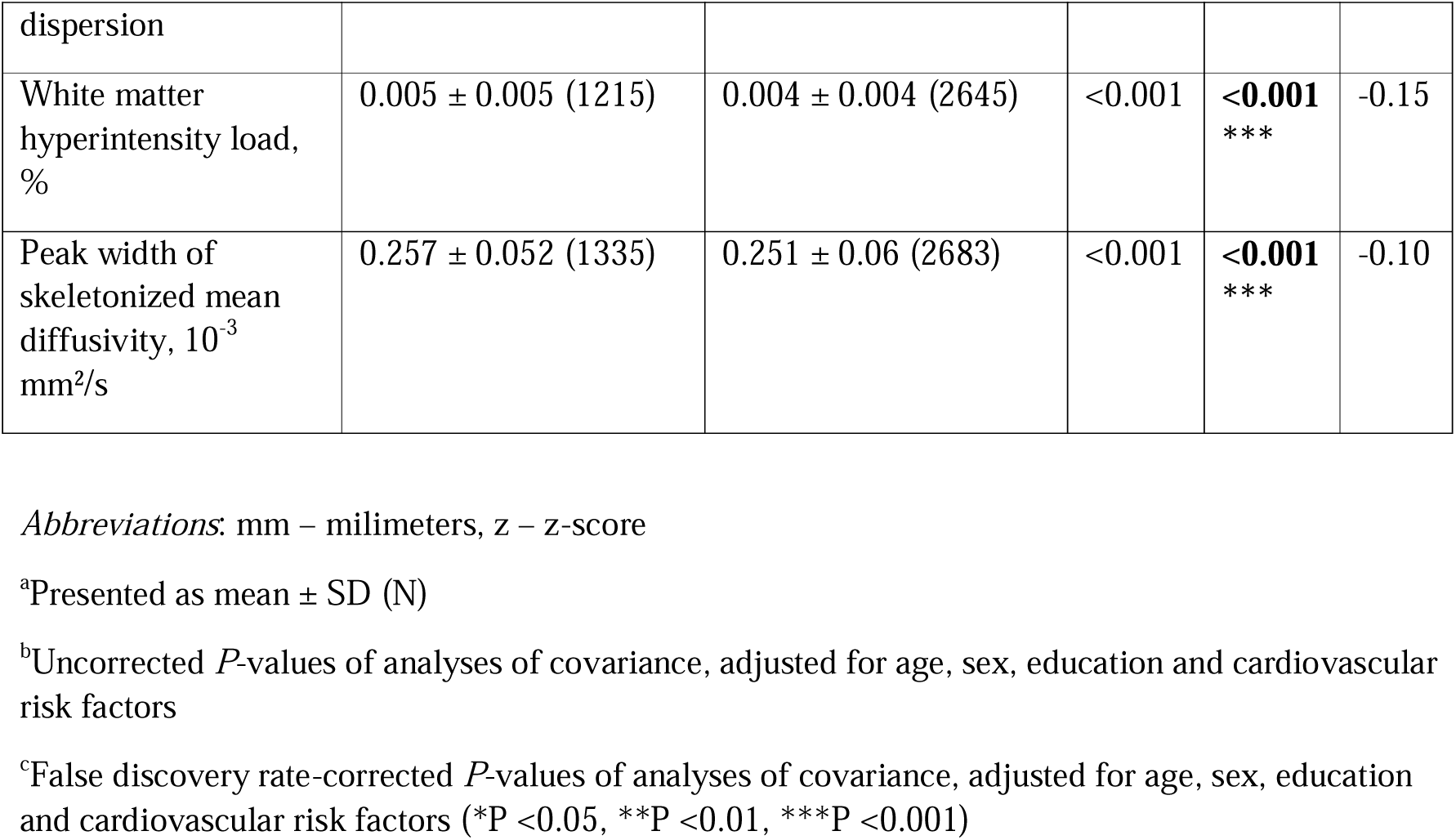
Results of cognitive and imaging assessments of individuals with atrial fibrillation compared to matched controls.

### Global analysis of imaging markers

To test for group differences in imaging markers averaged across the entire cortical gray matter or white matter, we conducted ANCOVAs, adjusted for sex, age, education and cardiovascular risk factors (see *table 2*).

Regarding markers of cortical macrostructure, subjects with AF showed a lower cortical thickness and gray matter volume compared to the control group (mean ± SD, cortical thickness [mm]: 2.367 ± 0.087 vs. 2.376 ± 0.088, Cohen’s *d* = 0.10, *P_FDR_* = 0.004; gray matter volume: 0.458 ± 0.048 vs. 0.461 ± 0.047, Cohen’s *d* = 0.06, *P_FDR_* = 0.032). There were no significant differences between the groups concerning cortical folding geometry.

In our analysis of global cortical microstructure markers, the AF group showed a higher isotropic volume fraction (0.254 ± 0.041 vs. 0.253 ± 0.039, Cohen’s *d* = -0.03, *P_FDR_* = 0.008), suggesting a heightened presence of extracellular free-water within the cortical tissue. Other markers including FA, MD, orientation dispersion, and neurite density in the gray matter were comparable between the groups.

Our investigation of mean skeletonized markers of white matter microstructure revealed both a lower global FA and a higher MD in the AF group compared to the control group (mean ± SD, FA: 0.408 ± 0.016 vs. 0.409 ± 0.018, Cohen’s *d* = 0.05, *P_FDR_* = 0.017; MD [10^-3^ mm^2^/s]: 0.810 ± 0.033 vs. 0.808 ± 0.33, Cohen’s *d* = -0.06, *P_FDR_* = 0.003). This reflects lower directionality of water diffusion with overall higher bulk diffusivity. Additionally, the AF group displayed a higher global isotropic volume fraction in the white matter (mean ± SD, 0.086 ± 0.014 vs. 0.085 ± 0.014, Cohen’s *d* = -0.08, *P_FDR_* = 0.008), indicating a higher proportion of extracellular free-water in the white matter. Global orientation dispersion and neurite density showed no significant group differences.

Evaluating white matter markers related to small vessel pathology, in subjects with AF we observed a higher WMH load (mean ± SD [%], 0.005 ± 0.005 vs. 0.004 ± 0.004, Cohen’s *d* = -0.15, *P_FDR_* < 0.001) and a higher PSMD (mean ± SD [10^-3^ mm^2^/s], 0.257 ± 0.052 vs. 0.251 ± 0.06, Cohen’s *d* = -0.10, *P_FDR_* < 0.001). These results remained stable when additionally controlling for AF comorbidities (*supplementary table S6)*.

### Mediation analysis

To investigate whether global imaging markers mediate the relationship between AF and cognitive domain performance, we performed a mediation analysis (*figure 2*).

**Figure 2.**
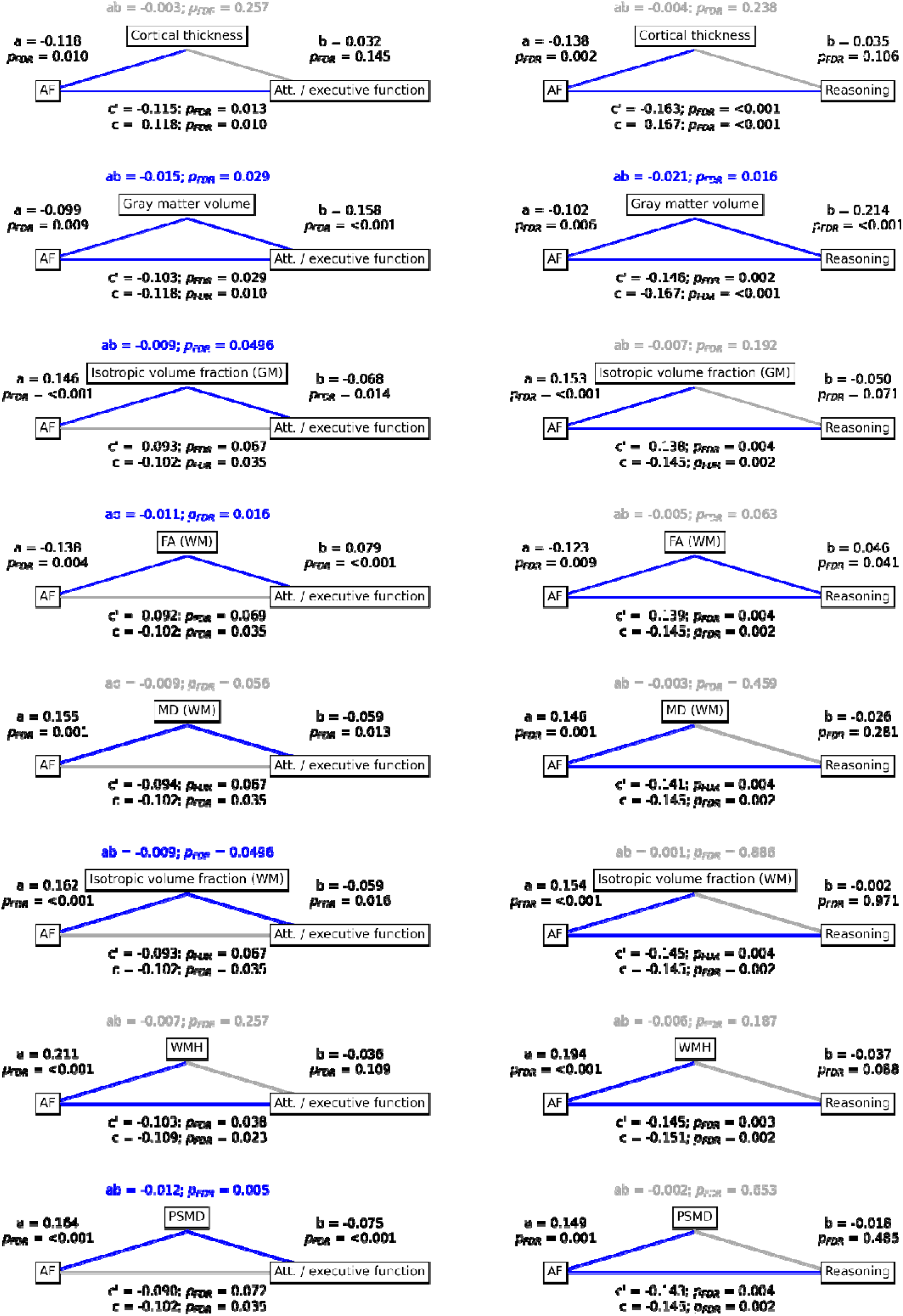
Mediation analysis. Mediation analysis results. Mediation effects of global imaging markers on the relationship between AF and attention/executive function as well as reasoning. Path plots display standardized effects and p-values: (a) AF to imaging marker, (b) imaging marker to cognitive score, (ab) indirect effect (c’) direct effect and (c) total effect. Significant paths are highlighted in blue; non-significant in light gray. If a relationship is significantly mediated, i.e., the indirect effect ab was significant and the direct effect c’ was reduced or non-significant compared to the total effect c, the text for ab is highlighted in blue. Only imaging markers with significant differences between AF and control groups are shown. The left path plots show results regarding attention/executive function, while the right plots depict those of reasoning. For path plots on all global imaging markers refer to *supplementary figure SX*. *Abbreviations*: AF = atrial fibrillation, FA = fractional anisotropy, GM = gray matter, MD = mean diffusivity, PSMD = peak width of skeletonized mean diffusivity, WM = white matter, WMH = white matter hyperintensity.

The relationship between AF and attention / executive function was found to be partially mediated by gray matter volume (ab = -0.015, *P_FDR_* = 0.029; c’ = -0.103, *P_FDR_* 0.029; c = -0.118, *P_FDR_* = 0.010) and fully mediated by gray matter isotropic volume fraction (ab = -0.009, *P_FDR_* = 0.00496; c’ = -0.093, *P_FDR_* 0.067; c = -0.102, *P_FDR_* = 0.035), white matter FA (ab = -0.011, *P_FDR_* = 0.016; c’ = -0.092, *P_FDR_* 0.069; c = -0.102, *P_FDR_* = 0.035), white matter isotropic volume fraction (ab = -0.009, *P_FDR_* = 0.0496; c’ = -0.093, *P_FDR_* 0.067; c = -0.102, *P_FDR_* = 0.035) and PSMD (ab = -0.012, *P_FDR_* = 0.005; c’ = -0.090, *P_FDR_* 0.072; c = -0.102, *P_FDR_* = 0.035).

The relationship between AF and reasoning was partially mediated by gray matter volume (ab = -0.021, *P_FDR_* = 0.016; c’ = -0.146, *P_FDR_* 0.002; c = -0.167, *P_FDR_* <0.001) and white matter FA (ab = -0.005, *P_FDR_* = 0.063; c’ = -0.139, *P_FDR_* 0.004; c = -0.145, *P_FDR_* = 0.002). For path plots showing mediation analysis results of all global imaging markers refer to *supplementary figures S8.* Scatter plots of the relationship between imaging markers and cognitive measures are shown in *supplementary figures S9-S11*.

### Regional distribution of cortical differences in macro- and microstructure

To detect spatial patterns of brain structural alterations, we performed ROI-wise analyses of gray and white matter imaging markers and TBSS.

Comparisons of Schaefer400-parcellated cortical thickness and gray matter volume revealed significant differences between the AF group and matched controls (*figure 2*). Individuals with AF exhibited lower cortical thickness in the primary sensorimotor and visual cortices, i.e., M1, S1, V1 as well as orbitofrontal and lateral prefrontal areas, the posterior insula, the superior temporal gyrus, bilaterally. Furthermore, individuals with AF showed lower bilateral gray matter volume in M1, S1, V1, as well as the middle and inferior temporal gyrus. Variations in cortical folding metrics were either restricted to specific parcels or were non-significant.

Comparisons of cortical microstructural measures revealed significant differences between the AF group and matched controls in the bilateral anterior cingulate, insula, medial prefrontal cortex, medial temporal lobe and precuneus (*figure 3*). These regions exhibited higher isotropic volume fraction and to varying degrees a lower FA and higher MD and orientation dispersion. Localized lower neurite density was found in in the anterior insular and adjacent zones. Cumulatively, the results identify microstructural irregularities in the affected areas, with alterations in the extracellular compartment. For statistical comparisons of all cortical effect maps please refer to *supplementary figure S12*.

**Figure 3.**
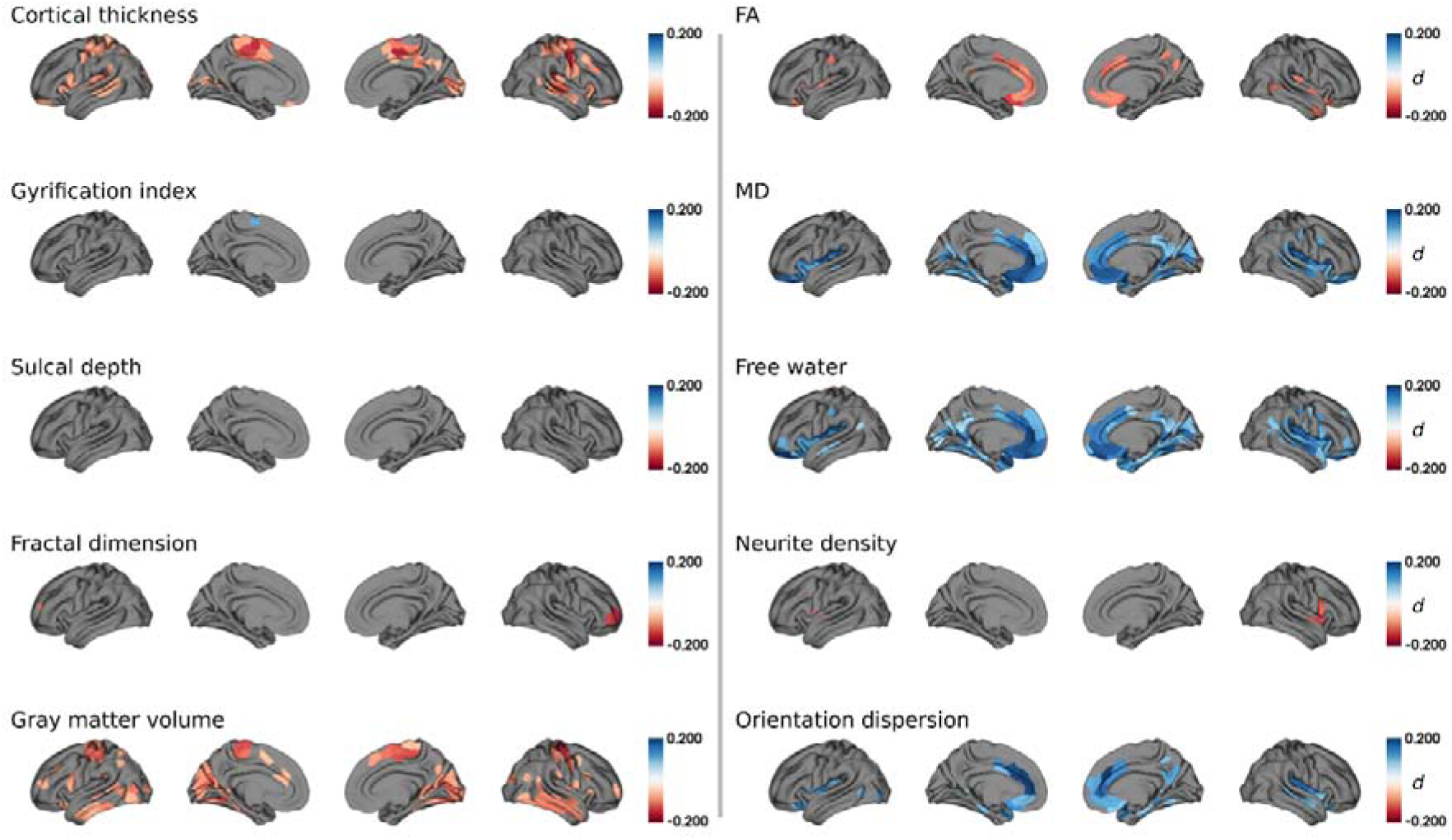
Group comparison of cortical macro- and microstructural indices. Regions of interest that significantly differed between groups are highlighted by colors encoding Cohen’s *d*: AF individuals < matched controls, red; AF individuals > matched controls, blue. *Abbreviations*: *d* = Cohen’s *d*, FA = fractional anisotropy, MD = mean diffusivity.

### Regional distribution of structural differences in the white matter

Analysis of microstructural measures in predefined anatomical white matter tracts disclosed widespread differences for FA, MD, isotropic volume fraction and orientation dispersion between the AF group and matched controls. Differences were found in multiple association, commissural and projection tracts, but only few brainstem and cerebellar tracts (corresponding details on all tracts investigated are illustrated in *figure 4* and *supplementary figures S13 – S16*): higher isotropic volume fraction in 47 (67.1%) tracts, lower FA was found in 43 (61.4%) tracts, higher MD in 47 (67.1%) tracts and higher orientation dispersion in 25 (35.7%) tracts. There were no significant differences for tract-level neurite density. Taken together, this imaging profile indicates extensive microstructural abnormalities throughout the white matter, with an emphasis on changes in the extracellular compartment.

**Figure 4.**
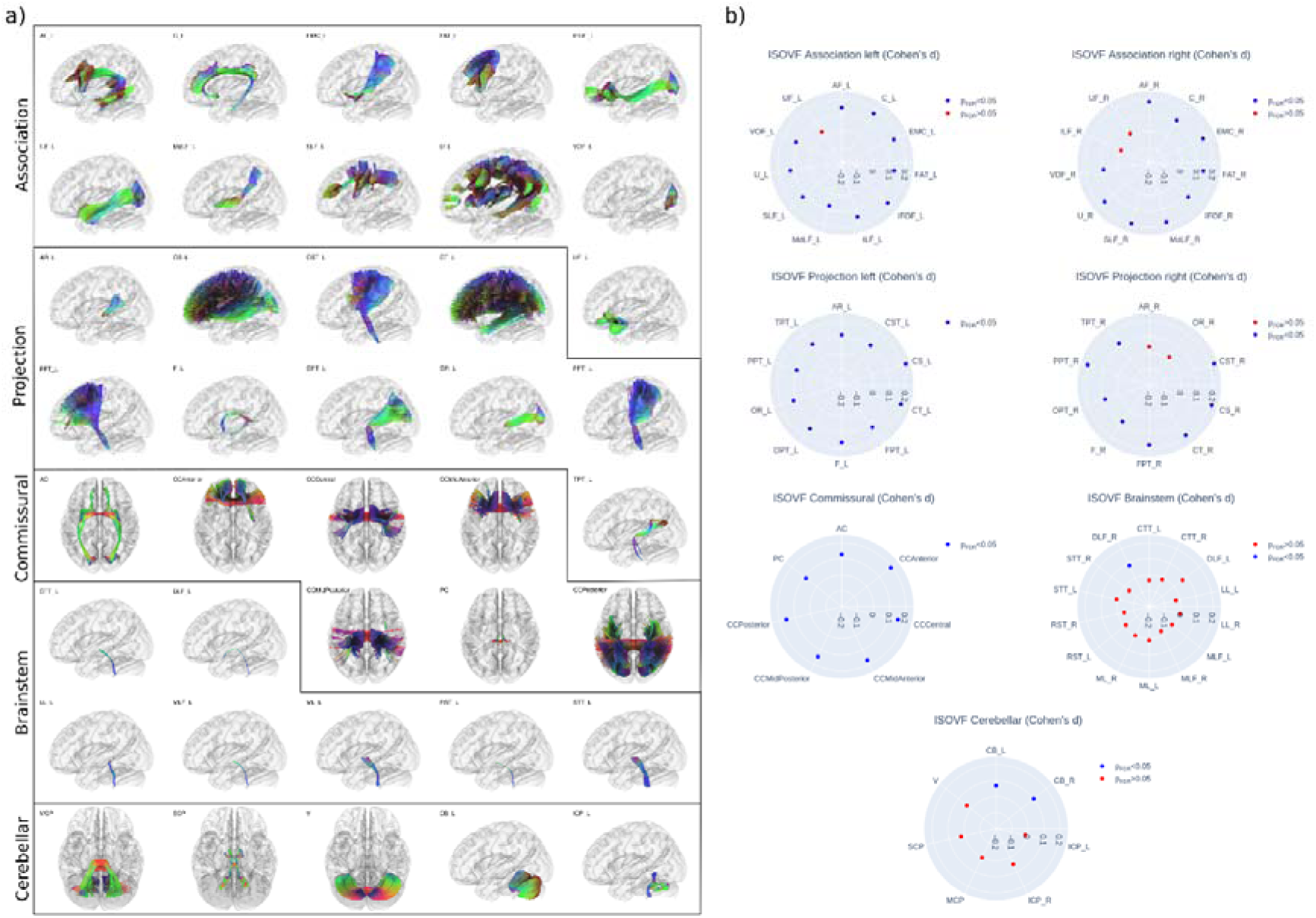
Group comparison of tract-level isotropic volume fraction. Left panel: anatomical depiction of the white matter tracts investigated, categorized into association, projection, commissural, brainstem, and cerebellar tracts. For paired tracts only left side examples are visualized. Right panel: radar plots represent the group differences (Cohen’s *d*) for isotropic volume fraction in each tract, with blue dots indicating significant differences and red dots marking non-significant differences. Each radar plot corresponds to one of the tract groups highlighted in the left panel. Tract abbreviations: Association tracts - AF = arcuate fascicle, C = cingulate, EMC = extreme capsule, FAT = frontal aslant tract, IFOF = inferior fronto-occipital fasciculus, ILF = inferior longitudinal fasciculus, MdLF = middle longitudinal fasciculus, SLF = superior longitudinal fasciculus, U = U-fibers, VOF = vertical occipital fasciculus, UF = uncinate fasciculus; Projection tracts – AR = acoustic radiation, CS = corticostriatal pathway, CST = corticospinal tract, CT = corticothalamic pathway, FPT = frontopontine tract, F = fornix, OPT = occipitopontine tract, OR = optic radiation, PPT = parietopontine tract, TPT = temporopontine tract; Commisural tracts – AC = anterior commissure, CC = corpus callosum, PC = posterior commissure; Brainstem tracts – CTT = central tegmental tract, DLF = dorsal longitudinal fasciculus, LL = lateral lemniscus, MLF = medial longitudinal fasciculus, ML = medial lemniscus, RST = rubrospinal tract, STT = spinothalamic tract; Cerebellar tracts – MCP = middle cerebellar peduncle, SCP = superior cerebellar peduncle, V = vermis, CB = cerebellum, ICP = inferior cerebellar peduncle.

We complemented the tract-level approach by TBSS, i.e., voxel-wise statistics on the entire white-matter skeleton. Corresponding results echoed tract-level findings by demonstrating widespread lower FA as well as higher MD and isotropic volume fraction in the white matter skeleton of AF individuals encompassing all brain lobes (*figure 5*). Group differences of neurite density and orientation dispersion were non-significant or spatially limited to few isolated voxel groups, respectively.

**Figure 5.**
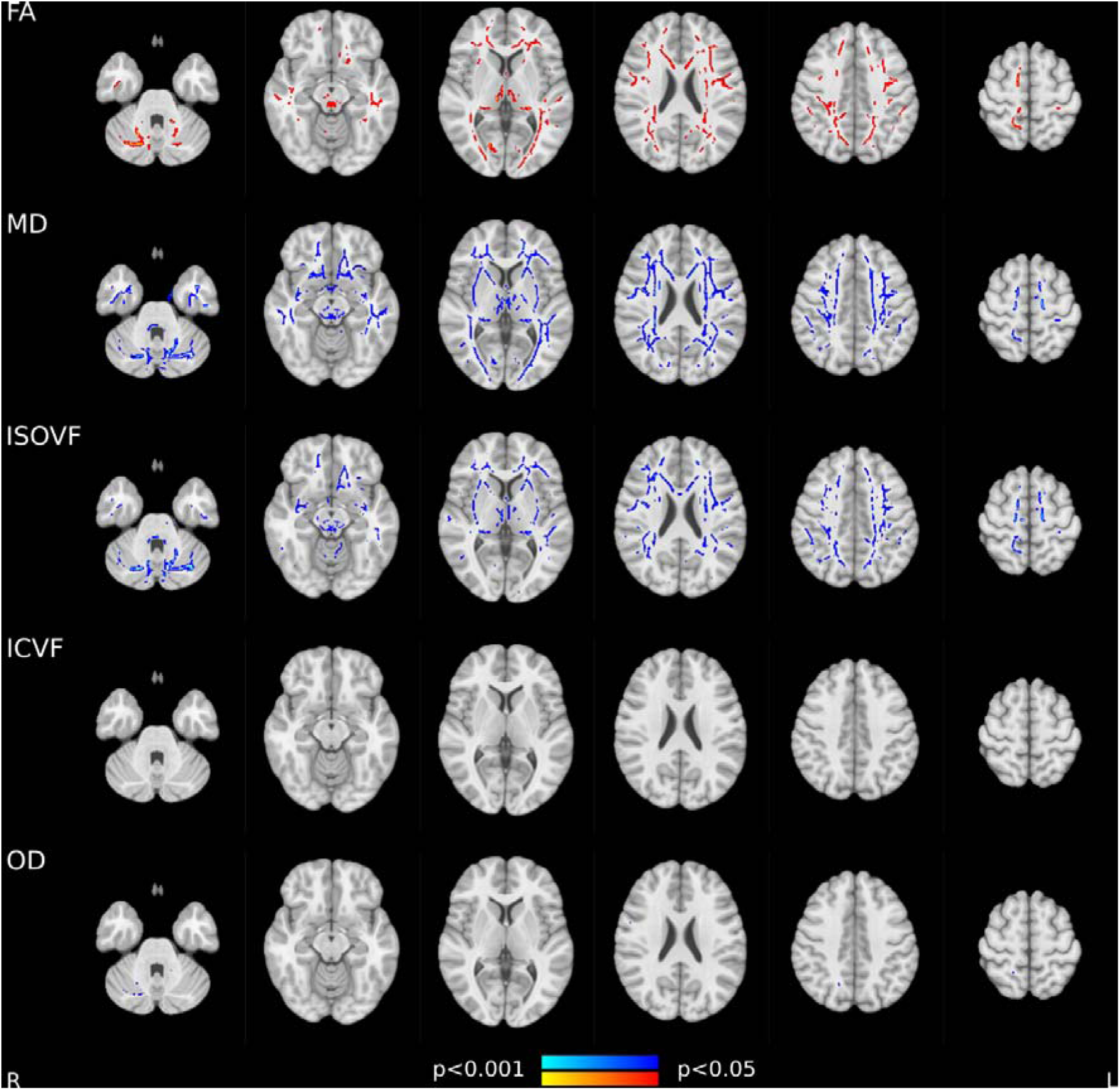
Tract-based spatial statistics of white matter imaging markers. Skeleton voxels that significantly differed between groups are highlighted by colors: AF individuals < matched controls, red; AF individuals > matched controls, blue. *Abbreviations*: FA = fractional anisotropy, MD = mean diffusivity.

## Discussion

In a cohort of individuals with AF without prior stroke or known dementia, we conducted an in-depth examination of cross-domain cognitive performance and multi-modal neuroimaging of brain macro- and microstructure. Our analysis revealed lower performance in the cognitive domains of attention / executive function and reasoning in individuals with AF. These cognitive deficits were accompanied by three main imaging findings: AF was associated with (1) macrostructural abnormalities of the cerebral cortex, including lower cortical thickness and gray matter volume predominantly in M1, S1, V1 as well as orbitofrontal, lateral prefrontal, posterior insular and temporal cortices; (2) microstructural cortical abnormalities of higher isotropic volume fraction in the anterior cingulate, insula, medial prefrontal cortex, medial temporal lobe and precuneus; and (3) widespread microstructural anomalies in the cerebral white matter, marked by lower FA as well as higher MD and isotropic volume fraction and higher burden of the small vessel disease markers WMH load and PSMD. Crucially, imaging markers statistically mediated the relationship of AF and cognitive domain performance. Overall, our findings identify a complex profile of structural brain changes that may provide novel links between AF and cognitive performance.

### AF is independently linked to cognitive deficits

Numerous prospective studies have observed an association between AF and cognitive disorders, spanning mild cognitive impairment to Alzheimer’s and vascular dementia.^39–41^ We examined the cognitive performance differences linked to AF in a population-based sample of stroke- and dementia-free. To isolate the cognitive effects attributable to AF, we matched relevant confounders and statistically adjusted for them and excluded subjects with prevalent neurological conditions. We observed a lower performance in cognitive domain scores of attention / executive function as well as reasoning implying a significant association of AF and cognitive impairment independent of confounders. These results remained robust when additionally controlling for the prevalence of common AF comorbidities including atherosclerotic heart disease and congestive heart failure. There were no differences in domain scores of information processing speed and memory performance; however, on individual cognitive test level, the lower performance in the Symbol Digit Substitution Test indicates reduced speed of information processing. Overall, effect sizes were modest, which might be attributed to our analytical design reducing cofounding effects, and the relatively senior age interval of our cohort. In previous studies cognitive effects of AF were stronger in individuals younger than 60 years.^42^ Our observations highlight deficits particularly in executive function while memory was not affected, echoing findings from previous population-based studies.^43–45^ Other studies reported associations across different cognitive domains, which led to the notion that AF impairs all major cognitive domains.^6^ We speculate that the differential associations of AF and cognition depend on stage-specific brain structural changes. Specifically, initial localized variations in tissue macro- and microstructure may be linked to focal cognitive deficits. As the disease advances, this pathology might spread to other regions, leading to widespread cognitive decline across multiple domains. Taken together, our results corroborate previous analyses reporting cognitive deficits in AF.

### AF is associated with altered cortical morphology

AF is considered to compromise brain health via complex, interacting effects on cerebral vasculature and parenchyma.^7^ To delineate these impacts, we investigated advanced neuroimaging markers of both gray and white matter integrity. Our results revealed abnormalities in both macro- and microstructural markers within the AF cohort.

We found altered cortical thickness and gray matter volume in AF individuals indicating neurodegenerative effects. AF might cause a decline in cortical thickness and gray matter volume by driving multiple vascular and inflammatory mechanisms, which impede proper blood flow and oxygenation, thus accelerating neuronal tissue loss. Although our study is the first to investigate markers of cortical macrostructure in AF, a negative correlation between AF and overall brain volume has been highlighted in previous reports, reinforcing our observations.^6,46–49^ On a regional scale, we found gray matter macrostructural differences to be localized bilaterally to primary sensorimotor and visual areas, orbitofrontal and lateral prefrontal cortices, the posterior insula and the temporal lobe. We interpret the relative symmetry of effects as indicative of vascular contributions, given the topological symmetry of vascular supply. Notably, the observed regions coincide with effect maps of prior research on cortical macrostructural effects of small vessel pathology in mild vascular cognitive impairment as well as in stroke.^50–52^ Thus, we hypothesize that the AF-induced injury of the cerebral cortex is relevantly influenced by small vessel disease, which in turn can lead to vascular cognitive deficits, offering a plausible connection between AF and cognitive decline, as also discussed for other cardiac diseases.^53^

### AF relates to extracellular free-water increases in gray and white matter

Turning to MRI markers of microstructural integrity, the AF group prominently displayed elevated isotropic volume fraction in the gray and white matter, indicative of an increase in extracellular free-water. Regional alterations in isotropic volume fraction were accompanied by changes in FA, denoting decreased water diffusion directionality; MD, indicating heightened overall water diffusivity; and orientation dispersion; implying fiber configuration shifts. Importantly, the neurite density index, a marker of cellular tissue, remained largely unchanged, suggesting preserved cellular and neurite integrity. These findings expand on a previous analysis showing globally reduced directionality of water diffusion in the white matter of individuals with AF.^49^ Taken together, the observed alterations in the different parameters derived from DWI likely reflect increased amounts of extracellular free-water alongside subtle cellular abnormalities like demyelination and dispersion of neurite orientations. We hypothesize that higher extracellular free-water and neurite dispersion in AF could stem from blood-brain barrier leakage and inflammation-driven osmotic shifts of water from blood to the extracellular space.

Our findings further highlight elevated global markers of small vessel pathology, specifically WMH load and PSMD, in subjects with AF. Hence, we argue that the microstructural effects in gray and white matter are relevantly attributable to AF-related small vessel pathology. This hypothesis aligns with previous reports indicating that extracellular free-water, as we observed in AF, is the primary contributor to tissue diffusion changes in small vessel pathology.^54,55^ Another study in a memory clinic cohort found that tissue free-water fully mediated the relationship between cardiac biomarkers and a five-year longitudinal cognitive decline, positioning the biomarker as a center piece for characterizing heart-brain interactions.^56^

### Brain structural differences mediate the link of AF and cognition

Integrating the observed group differences in cognitive and imaging markers, we conducted a mediation analysis to statistically assess brain structure as a potential pathomechanistic link between AF and cognitive function. We found that the association between AF and cognitive function was significantly mediated by structural imaging markers. Specifically, the relationship between AF and attention/executive function was mediated by gray matter volume, gray matter isotropic volume fraction, white matter FA, white matter isotropic volume fraction, and PSMD and between AF and reasoning by gray matter volume and white matter FA. These results suggest that AF’s link to cognitive function depends to a relevant extent to differences in these imaging markers, underscoring the significance of macrostructural and microstructural brain changes in the cognitive impact of AF.

### Microstructural abnormalities map to specific cortical networks

Our study pinpointed AF-related variations in gray matter microstructure, with symmetrically impacted regions including the anterior cingulate, insula, medial prefrontal cortex, medial temporal lobe and precuneus. This observation becomes especially notable when contextualized with two pathomechanistic theories explaining its origin.

First, the pattern might be a manifestation of advancing AF-related cerebral pathophysiology. The anterior cingulate, medial temporal lobe and precuneus have a particular role in Alzheimer’s and vascular dementia, directly influencing associations with cognitive symptoms.^57–60^ It is plausible that pathology in these areas might accelerate cognitive decline and the onset of dementia in individuals with AF. As there is an observable disparity between cortical patterns of macro- and microstructural changes, we propose that these differences could represent varying local stages of tissue changes all tethered to a shared disease mechanism. An increase in extracellular free-water might act as a precursor to more pronounced neurodegenerative processes. Thus, the regions showing microstructural deviations in our current cross-sectional analysis might manifest macrostructural changes as the disease progresses. This interpretation finds support in studies assessing cortical thickness in diverse stages of vascular cognitive impairment. These studies demonstrate that thickness differences between mild and severe cases of vascular cognitive impairment coincide with the cortical regions highlighted by altered microstructure in our analysis which might be due to common disease mechanisms.^50,51^ Further support comes from observations of microstructural indices being demonstrably highly sensitive to subtle disease alterations in vascular cognitive impairment and preceding morphological effects and lesion occurence.^61,62^

Second, on a more speculative note, the observed associative effects could also be attributed to neurodegenerative processes increasing the risk of AF. The regions we identified are all components of the central autonomic network (CAN), which plays a pivotal role in regulating heart rate and contractility by transmitting autonomic signals.^63,64^ Chronic ischemic and degenerative changes within the central autonomic network, possibly associated with CSVD, might heighten the propensity for AF development. This perspective is supported by preclinical studies, which have demonstrated neurogenically triggered cardiac arrhythmias and changes in the left atrial myocardium post-stroke in rodent models.^65,66^ Furthermore, in cases of AF detected after stroke (AFDAS), stroke lesions predominantly appear within the central autonomic network, pointing towards a potential neurogenic origin.^63^ Notably, lesions in the right anterior insula correlate with post-stroke acute myocardial injury, underscoring the region’s significance in brain-heart interactions.^67^ In fact, the anterior insula is the only area showing significant group differences of neurite density in our analysis, implying greater intracellular volume loss possibly due to cellular and axonal damage. Longitudinal neuroimaging studies are needed to further substantiate these hypotheses in AF.

### A unifying hypothesis to explain brain imaging correlates of AF in the normal population

Drawing upon a comprehensive analysis of brain imaging correlates associated with AF, our research converges on a unifying hypothesis: small vessel pathology stands out as a key mechanism connecting AF to cognitive decline in individuals unaffected by stroke or dementia. This assertion is supported by several key observations. Firstly, our results underscore the involvement of executive dysfunction and information processing speed both being key cognitive correlates of small vessel disease, while memory was not affected in our cohort of AF subjects.^68^ Secondly, the observed macro- and microstructural differences in the cortex echo patterns reflecting various progression stages of vascular cognitive impairment. Lastly, the widespread alterations in the white matter mirror a neuroimaging profile consistent with small vessel disease, accompanied by heightened WMH load and PSMD.^69^

Our findings not only shed light on the intricate relationships between AF, cognitive decline, and neuroanatomical changes, but they also hint at potential avenues of clinical utilization. The definitive role of AF treatments, particularly in precluding cognitive comorbidities, is yet to be firmly established. However, there is evidence suggesting that interventions targeting AF might reverse prothrombogenic and proinflammatory cascades, as well as recovering cerebral perfusion.^70^ Notably, there have been promising outcomes related to oral anticoagulation, pointing towards enhanced cognitive trajectories.^71,72^ Preliminary observational datasets also hint at the potential cognitive benefits associated with rhythm control strategies, either pharmacological or non-pharmacological.^73^ Moving forward, leveraging neuroimaging could enable patient-tailored therapeutic interventions, facilitating to identify subgroups at risk of cognitive disorders likely to reap the most substantial benefits.

### Strengths and limitations

A strength of our study is the utilization of large-scale clinical and multimodal MRI data, offering a detailed view of the link of AF and brain health, supported the use of advanced neuroimaging methods. Nonetheless, we acknowledge limitations of our work. First, due to the indirect nature of the performed imaging-phenotype association analyses, our findings cannot firmly establish causal links and should be regarded as hypothesis-generating. Second, despite controlling for cardiovascular risk factors through matching and assessing the effects of comorbidities in our statistical models, we cannot entirely rule out the influence of AF comorbidities on the relationship between AF, brain structure, and cognition. Third, our diagnosis of AF, based on ICD codes from the UK Biobank instead of electrocardiography, may pose potential classification concerns. Lastly, by not distinguishing between AF subtypes such as permanent or paroxysmal, we might overlook subtype-specific associations. Taken together, longitudinal and experimental studies are needed that expand on our findings to further discern AF effects on brain health.

## Conclusion

Our comprehensive exploration of a large cohort of individuals with AF unveiled clear links between AF, structural brain changes and cognitive decline, notably in individuals without a history of stroke or dementia. Our research emphasizes small vessel pathology as a potential mechanism associating AF with cognitive deficits. We observed cognitive deficits and significant macro- and microstructural brain alterations mirroring those observed in vascular cognitive impairment. As this research field progresses, harnessing neuroimaging could pave the way for individualized therapeutic strategies.

## Acknowledgments

The authors wish to acknowledge all participants of the UK Biobank. This research has been conducted using the UK Biobank Resource under Application Number 71359.

## Author Contributions

We describe contributions to the paper using the CRediT contributor role taxonomy. M.P.: Conceptualization, Data Curation, Formal analysis, Investigation, Methodology, Project administration, Resources, Software, Visualization, Writing—original draft, Writing—review & editing; C.C.: Writing—review & editing; F.L.N.: Software, Writing—review & editing; T.I.: Writing—review & editing; R.S.: Writing—review & editing; A.O.: Writing—review & editing; F.H.: Writing—review & editing; K.P.: Writing—review & editing; S.B.E.: Funding acquisition, Writing—review & editing; P.K.: Writing—review & editing; E.S.: Writing—review & editing; B.C.: Resources, Funding acquisition, Writing—review & editing; G.T.: Conceptualization, Funding acquisition, Supervision, Writing—review & editing; M.J.: Conceptualization, Supervision, Writing—original draft, Writing—review & editing.

