## Supplementary materials for "Mapping the Interplay of Atrial Fibrillation, Brain Structure and Cognitive Dysfunction"

–

*Supporting Information*


Marvin Petersen, MD[a]; Céleste Chevalier, MD[b,c]; Felix L. Naegele, MD[a]; Thies Ingwersen, MD[a]; Amir Omidvarnia[d,e], PhD; Felix Hoffstaedter[d,e], PhD; Kaustubh Patil[d,e], PhD; Simon B. Eickhoff[d,e], MD; Renate B. Schnabel, MD[b,c]; Paulus Kirchhof[b,c], M.D; Eckhard Schlemm, MBBS, PhD[a]; Bastian Cheng, MD[a]; Götz Thomalla, MD[a]; Märit Jensen, MD[a]

[a]Department of Neurology, University Medical Center Hamburg-Eppendorf, Hamburg, Germany

[b]Department of Cardiology, University Heart and Vascular Center, Hamburg, Germany

[c]DZHK (German Center for Cardiovascular Research), partner site Hamburg/Kiel/Luebeck, Germany

[d]Institute for Systems Neuroscience, Medical Faculty, Heinrich-Heine University Düsseldorf, Düsseldorf, Germany

[e]Institute of Neuroscience and Medicine, Brain and Behaviour (INM-7), Research Center Jülich, Jülich, Germany

#### Content

Methods ... 3
- Table S1 – Excluded diagnoses based on non-cancer illness information ... 3
- Text S2 - Image processing ... 4
  - T1-weighted MRI ... 4
  - Diffusion-weighted MRI ... 5
  - Fluid-attenuated inversion recovery MRI ... 7
  - Quality assurance ... 7

Results ... 8
- Figure S3 - Sample selection flowchart ... 8
- Figure S4 – Matching results visualized as balance plot ... 9
- Figure S5 – AF comorbidities ... 10
- Figure S6 – Group comparisons controlling for AF comorbidities ... 11
- Figure S7 – Group comparisons of individual cognitive tests ... 13
- Figure S8 – Mediation analysis results ... 14
- Figure S9 – Regression analysis: cognition ~ cortical macrostructural imaging markers ... 16
- Figure S10 – Regression analysis: cognition ~ cortical microstructural imaging markers ... 17
- Figure S11 – Regression analysis: cognition ~ white matter imaging markers ... 18
- Figure S12 – Cortical effect maps comparison ... 19
- Figure S13 - Group comparison of tract-level fractional anisotropy (FA) ... 20
- Figure S14 - Group comparison of tract-level mean diffusivity (MD) ... 21
- Figure S15 - Group comparison of tract-level neurite density index ... 22
- Figure S16 - Group comparison of tract-level orientation dispersion ... 23

References ... 24

#### Methods

##### Table S1 – Excluded diagnoses based on non-cancer illness information

| ID | Diagnosis |
|---|---|
| 1081 | stroke |
| 1082 | transient ischaemic attack (tia) |
| 1083 | subdural haemorrhage/haematoma |
| 1086 | subarachnoid haemorrhage |
| 1240 | neurological injury/trauma |
| 1244 | infection of nervous system |
| 1245 | brain abscess/intracranial abscess |
| 1246 | encephalitis |
| 1247 | meningitis |
| 1258 | chronic/degenerative neurological problem |
| 1261 | multiple sclerosis |
| 1262 | parkinsons disease |
| 1263 | dementia/alzheimers/cognitive impairment |
| 1264 | epilepsy |
| 1266 | head injury |
| 1408 | alcohol dependency |
| 1409 | opioid dependency |
| 1410 | other substance abuse/dependency |
| 1434 | other neurological problem |
| 1491 | brain haemorrhage |
| 1583 | ischaemic stroke |
| 1626 | fracture skull / head |

#### Text S2 - Image processing

##### T1-weighted MRI

T1w image processing was performed in CAT12.[1] CAT12 processing can be divided into 2 streams: (1) surface-based morphometry (SBM), and (2) voxel-based morphometry (VBM). Together, SBM and VBM are imaging methods that enable the identification and characterization of macrostructural brain abnormalities.

###### *Surface-based morphometry*

SBM results in vertex-level measures of cortical thickness and cortical folding, i.e., point-wise estimates of cortical geometry. Surface-based morphometry was performed in two stages: (1) surface creation and registration and (2) computation of morphometric estimates. In the surface creation phase, CAT12 utilized a projection-based thickness method to derive the initial cortical thickness and central surface. Subsequently, the central surface underwent refinement and topological defect correction, yielding final central, pial, and white matter surface meshes. These refined surfaces were then employed to re-estimate cortical thickness using the FreeSurfer thickness method, which assesses the width of the gray matter ribbon by measuring the distance between its inner and outer boundaries.[2] Furthermore, the final central surface served for the computation of three cortical folding metrics. The gyrification index was computed as the "smoothed absolute mean curvature", which involved averaging curvature values from each vertex of a spherical surface mesh within a 3 mm radius.[3] Larger negative gyrification index values signify sulci, while positive values represent gyri. Sulcal depth was computed as the distance between a point on the sulcal surface and the nearest point on the brain's convex hull, which envelopes the cortex's outermost (pial) surface.[4] Deeper sulcal depths indicate more pronounced sulci, while shallower depths suggest less pronounced ones. Fractal dimension was calculated as the slope of a logarithmic plot comparing surface area to the maximum l-value of the surface reconstruction (a measure of the bandwidth used to reconstruct the surface shape), with a higher FD indicating greater complexity and irregularity of cortical folding, and a lower FD denoting a smoother surface.[5] Cortical thickness was smoothed using a 12mm kernel, while the cortical folding metrics were smoothed with a 20mm kernel.

##### *Voxel-based morphometry*

Voxel-based morphometry (VBM) results in voxel-wise estimates of gray matter volume. In CAT12, the VBM protocol include a tissue segmentation and spatial registration procedure. First, the T1w was denoised by applying a spatially adaptive non-local means (SANLM) filter. After the denoising, SPM's unified segmentation was performed, producing segmentation maps of multiple brain tissues, including a gray matter map. These gray matter segmentations capture the voxel-wise gray matter content. The tissue segmentations were registered to standardized MNI space templates using Geodesic Shooting.[6] The registration information was then incorporated into the gray matter segmentation by multiplying the deformation maps with the gray matter segmentation maps in a "modulation step." This adjustment accounts for volume alterations during spatial normalization, thereby estimating local gray matter volume in native space.[7] Finally, modulated gray matter volume maps were smoothed using a 6mm Gaussian kernel.

#### Diffusion-weighted MRI

##### *Preprocessing*

Analysis of diffusion-weighted MR images (DWI) was based on data already preprocessed by the UK Biobank. Details on UK Biobank preprocessing procedures can be found online (https://biobank.ctsu.ox.ac.uk/crystal/crystal/docs/brain_mri.pdf). In brief, DWI preprocessing included correction of eddy currents, head motion and outlier-slice removal using FSL's eddy as well as gradient distortion correction.[8]

##### *Diffusion tensor imaging and Neurite Orientation Dispersion and Density Imaging*

Fractional anisotropy (FA) and mean diffusivity (MD) were derived from diffusion tensors which were modelled based on preprocessed DWI using a least-squares fit.[9,10] FA and MD provide insights into the directionality and magnitude of water diffusion within tissues, respectively. In addition, Neurite Orientation Dispersion and Density Imaging (NODDI) was performed using

Accelerated Microstructure Imaging via Convex Optimization (AMICO, https://github.com/daducci/AMICO).[11,12] NODDI offers an assessment of the brain's microstructure beyond conventional DTI markers by capitalizing on a multi-compartment model. This model dissects the intricate features of neural tissue into distinct compartments. (1) The intracellular compartment, which showcases hindered or restricted diffusion, is quantified using the neurite density index (also intracellular volume fraction, ICVF). This measure essentially captures the proportion of space occupied by neuronal and glial cell bodies and their processes within a given voxel. (2) The extracellular compartment is characterized by isotropic diffusion, indicating equal diffusion in all directions. This is represented by measurement of isotropic volume fraction (ISOVF), which reflects the volume occupied by the space outside the cellular structures. Neurite orientations, a key aspect of neural tissue microstructure and organization, are described by orientation dispersion. This metric provides insights into the variability in the alignment of axons and dendrites within a voxel. Together, DTI and NODDI measures provide a detailed understanding of the underlying tissue microarchitecture, aiding in deciphering the intricate complexities of neural structures.

##### *Tract-based spatial statistics*

In order to derive skeletonized maps of each of the estimated diffusion parameters, we conducted tract-based spatial statistics (*TBSS*) utilizing the standardized FA template from FSL as the registration target.[13,14] Put briefly, individual FA images in template space got eroded to exclude non-brain voxels on the outer edge of the image. Next, a valid mask containing only the intersection of all subjects' brains was derived and used to mask the average of all previously eroded FA images. This mean FA image was subsequently used to derive a white matter skeleton. Next, all individual FA images were projected onto the mean FA skeleton. The resultant projection vectors were used to skeletonize all of the remaining diffusion metrics, i.e. MD and NODDI markers.

##### *Peak width of skeletonized mean diffusivity*

Peak width of skeletonized mean diffusivity (PSMD) was calculated based on standard procedures.[15] PSMD was calculated as the difference between the 95th and 5th percentile of MD values on the white matter skeleton in standard (MNI) space. A mask supplied by the developers was used to exclude white matter areas susceptible to partial volume effects of cerebrospinal fluid (https://github.com/miac-research/psmd/blob/main/skeleton_mask_2019.nii.gz).

#### Fluid-attenuated inversion recovery MRI

##### *White matter hyperintensity segmentation*

Precomputed WMH segmentation masks of the UK Biobank were used. To obtain them, FSL's *Brain Intensity AbNormality Classification Algorithm* (*BIANCA*)[16] was applied on FLAIR images and T1w images for white matter hyperintensity (WMH) segmentation. The WMH load was calculated as the WMH volume divided by the total intracranial volume calculated by CAT12.

#### Quality assurance

In order to minimize the influence of noise and artifacts, quality assurance (QA) of MRI data was conducted both quantitatively and qualitatively. For raw and processed T1w data, an image quality rating and sample homogeneity score were derived with CAT12. Visual QA of T1w imaging data was subsequently performed for individuals with an IQR and sample homogeneity score <0.8. For DWI, visual QA was performed on raw DWI and skeletonized diffusion indices in individuals with mean skeletonized diffusion metrics ± 2.5 standard deviations from the mean.

### Results

#### Figure S3 - Sample selection flowchart

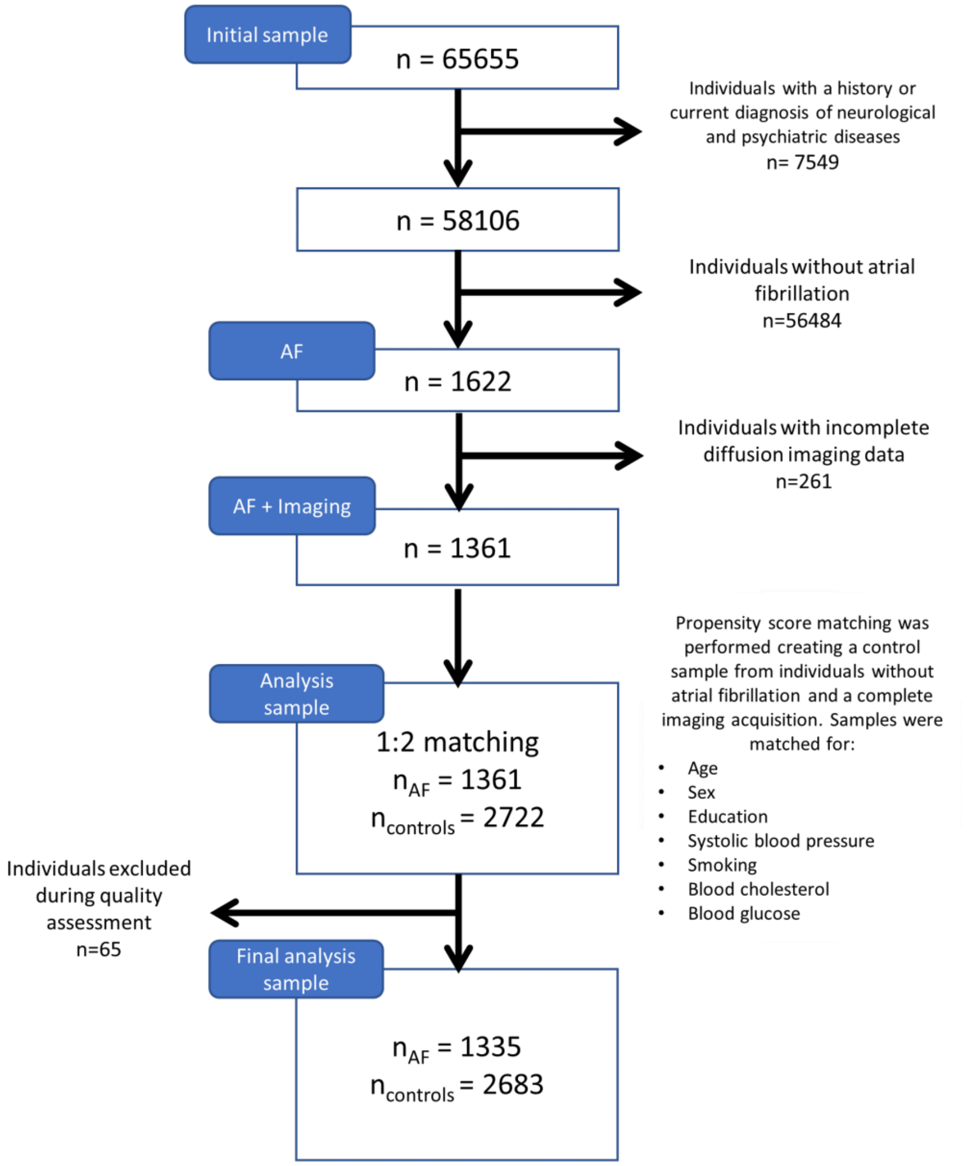

#### Figure S4 – Matching results visualized as balance plot

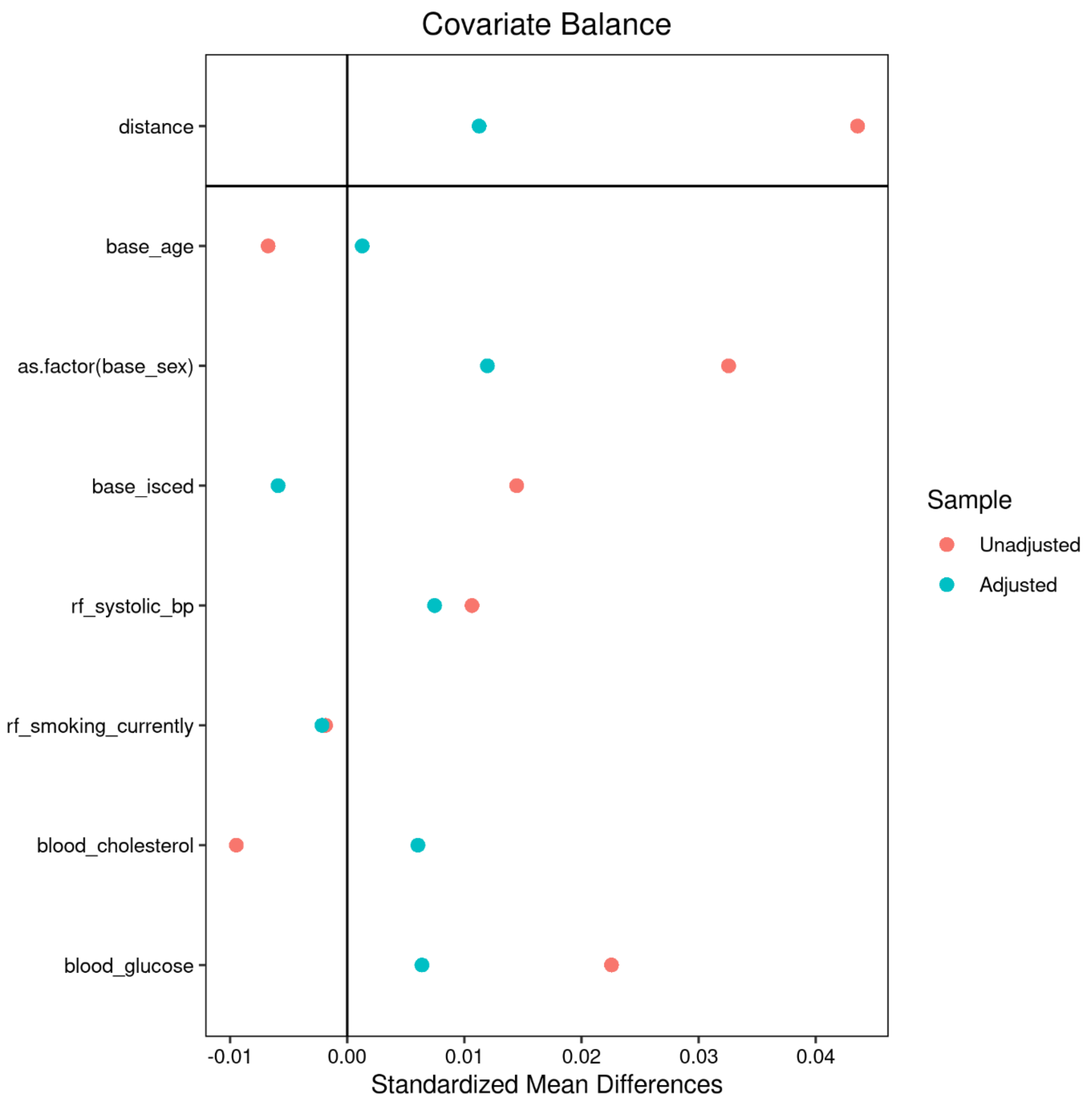


Standardized mean differences between the atrial fibrillation and healthy control group for each matching variable before (unadjusted, red) and after matching (adjusted, turquoise) are displayed. The closer the standardized mean difference is to zero, the more similar the groups are. Each matching variable is depicted separately, i.e., from top to bottom, age, sex, education, systolic blood pressure, smoking, blood cholesterol and blood glucose.

#### Figure S5 – AF comorbidities

| Diagnosis[a] | Atrial fibrillation | Matched controls | *P*[c] |
|---|---|---|---|
| **Atherosclerotic heart disease (I25.1)** | 253 (18.95) | 195 (7.26) | <0.001 |
| **Congestive heart failure (I50.0)** | 50 (3.74) | 4 (0.15) | <0.001 |
| **Hyperthyreoidism (E05.0, E05.1, E05.2)** | 3 (0.22) | 3 (0.11) | 0.660 |
| **Diabetes mellitus type II, (E10, E11)** | 120 (9.09) | 185 (6.96) | 0.023 |
| **Alcohol abuse (F10.1, F10.2)** | 23 (1.72) | 31 (1.16) | 0.184 |
| **Chronic obstructive sleep apnea (G47.3)** | 55 (4.12) | 50 (1.86) | <0.001 |

[a]ICD10 codes in parentheses
[b]Presented as n (%)
[c]uncorrected *P*-values of $\chi 2$ tests

#### Figure S6 – Group comparisons controlling for AF comorbidities

| Measure | Atrial fibrillation[a] | Matched controls[a] | $P_{uncorr}$[b] | $P_{FDR}$[c] | *Cohen's d* |
|---|---|---|---|---|---|
| **Cognitive domains** | | | | | |
| Attention / executive dysfunction, z | -0.058 ± 0.851 (787) | 0.039 ± 0.820 (1817) | 0.023 | 0.047* | 0.12 |
| Information processing speed, z | -0.039 ± 0.720 (836) | 0.031 ± 0.722 (1895) | 0.116 | 0.154 | 0.10 |
| Memory, z | -0.007 ± 0.570 (779) | 0.012 ± 0.556 (1749) | 0.710 | 0.710 | 0.03 |
| Reasoning, z | -0.085 ± 0.837 (843) | 0.049 ± 0.838 (1893) | 0.001 | 0.004* | 0.16 |
| **Gray matter** | | | | | |
| Cortical thickness, mm | 2.367 ± 0.087 (1293) | 2.376 ± 0.088 (2594) | 0.008 | 0.024* | 0.10 |
| Gyrification index | 27.623 ± 0.501 (1293) | 27.629 ± 0.517 (2594) | 0.770 | 0.815 | 0.01 |
| Sulcal depth, mm | 9.134 ± 0.471 (1293) | 9.135 ± 0.472 (2594) | 0.959 | 0.959 | 0.00 |
| Fractal dimension | 2.613 ± 0.024 (1293) | 2.614 ± 0.024 (2594) | 0.352 | 0.512 | 0.04 |
| Gray matter volume | 0.458 ± 0.048 (1293) | 0.461 ± 0.047 (2594) | 0.083 | 0.150 | 0.06 |
| Fractional anisotropy | 0.144 ± 0.093 (1335) | 0.144 ± 0.089 (2683) | 0.370 | 0.512 | -0.01 |
| Mean diffusivity, $10^{-3}$ mm²/s | 1.083 ± 0.080 (1335) | 1.083 ± 0.076 (2683) | 0.148 | 0.243 | -0.01 |
| Isotropic volume fraction | 0.254 ± 0.041 (1335) | 0.253 ± 0.039 (2683) | 0.025 | 0.057 | -0.03 |
| Neurite density index | 0.343 ± 0.024 (1335) | 0.342 ± 0.021 (2683) | 0.741 | 0.815 | -0.02 |
| Orientation dispersion | 0.455 ± 0.018 (1335) | 0.455 ± 0.017 (2683) | 0.541 | 0.649 | -0.00 |

| White matter | | | | | |
|---|---|---|---|---|---|
| Fractional anisotropy (FA) | 0.408 ± 0.016 (1335) | 0.409 ± 0.018 (2683) | 0.014 | 0.035* | 0.05 |
| Mean diffusivity (MD), $10^{-3}$ mm²/s | 0.810 ± 0.033 (1335) | 0.808 ± 0.33 (2683) | 0.003 | 0.013* | -0.06 |
| Isotropic volume fraction | 0.086 ± 0.014 (1335) | 0.085 ± 0.014 (2683) | <0.001 | 0.002** | -0.08 |
| Neurite density index | 0.554 ± 0.028 (1335) | 0.554 ± 0.027 (2683) | 0.458 | 0.589 | 0.00 |
| Orientation dispersion | 0.256 ± 0.011 (1335) | 0.255 ± 0.012 (2683) | 0.076 | 0.150 | -0.04 |
| White matter hyperintensity load, % | 0.005 ± 0.005 (1215) | 0.004 ± 0.004 (2645) | <0.001 | <0.001*** | -0.15 |
| Peak width of skeletonized mean diffusivity, 10-3 mm²/s | 0.257 ± 0.052 (1335) | 0.251 ± 0.06 (2683) | <0.001 | 0.002** | -0.10 |

Abbreviations: z – z-score
[a]Presented as mean ± SD (N)
[b]Uncorrected P values of analyses of covariance, adjusted for age, sex, education, cardiovascular risk factors and AF comorbidities
[c]False discovery rate-corrected P values of analyses of covariance, adjusted for age, sex, education, cardio-vascular risk factors and AF comorbidities (*P <0.05, **P <0.01, ***P <0.001)

#### Figure S7 – Group comparisons of individual cognitive tests

| Clinical measure[a] | Atrial fibrillation | Matched controls | $P_{uncorr}$[b] | $P_{FDR}$[c] | Cohen's d |
|---|---|---|---|---|---|
| **Attention and executive function** | | | | | |
| Tower Rearranging Test | 9.18 ± 3.24 (840) | 9.55 ± 3.15 (1891) | 0.008 | **0.034*** | 0.12 |
| Trail Making Test part B, seconds | 664.67 ± 309.68 (804) | 634.50 ± 295.77 (1855) | 0.023 | 0.058 | -0.10 |
| **Processing speed** | | | | | |
| Reaction Time Test | 6.71 ± 0.17 (1213) | 6.72 ± 0.17 (2453) | 0.139 | 0.231 | 0.04 |
| Symbol Digit Substitution Test | 16.68 ± 5.08 (848) | 17.28 ± 5.04 (1915) | 0.010 | **0.034*** | 0.12 |
| Trail Making Test part A, seconds | 250.38 ± 91.47 (851) | 248.11 ± 98.84 (1916) | 0.904 | 0.904 | -0.02 |
| **Memory** | | | | | |
| Numeric Memory Test | 6.48 ± 1.66 (883) | 6.55 ± 1.61 (1977) | 0.36 | 0.4 | 0.04 |
| Paired Associate Learning Test | 6.27 ± 2.68 (867) | 6.44 ± 2.70 (1941) | 0.221 | 0.316 | 0.06 |
| Prospective Memory Test | 1.08 ± 0.45 (1224) | 1.06 ± 0.45 (2476) | 0.256 | 0.319 | -0.04 |
| **Reasoning** | | | | | |
| Fluid Intelligence Test | 6.34 ± 2.02 (1192) | 6.49 ± 2.07 (2414) | 0.034 | 0.068 | 0.07 |
| Matrix Pattern Completion Test | 7.40 ± 2.07 (848) | 7.68 ± 2.07 (1911) | 0.003 | **0.027*** | 0.13 |

Abbreviations: z – z-score
[a]Presented as mean ± SD (N)
[b]Uncorrected P values of analyses of covariance, adjusted for age, sex and years of education
[c]False discovery rate-corrected P values of analyses of covariance, adjusted for age, sex, education and cardiovascular risk factors (*P <0.05, **P <0.01, ***P <0.001)

### Figure S8 – Mediation analysis results

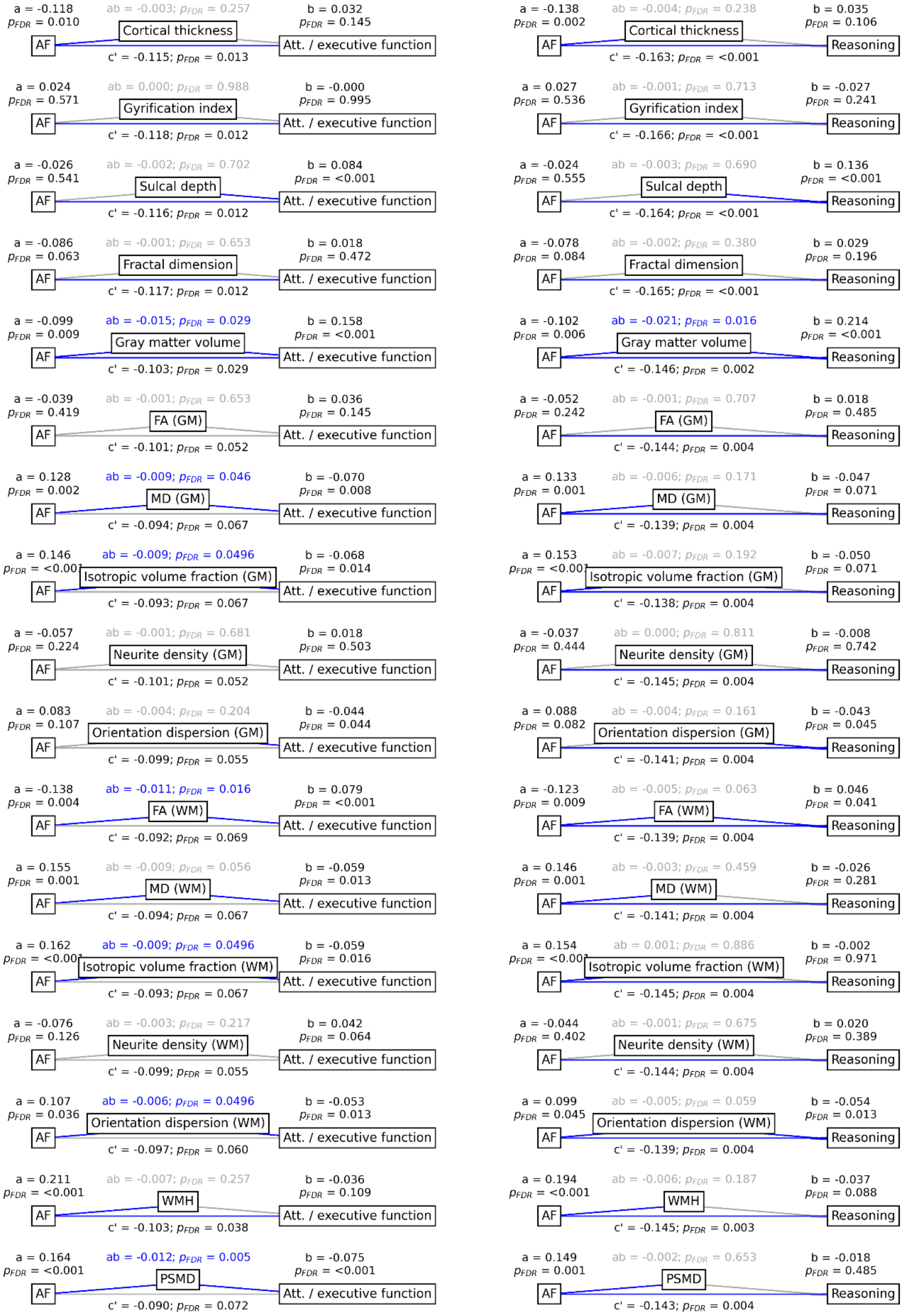

Mediation analysis results. Mediation effects of global imaging markers on the relationship between AF and attention/executive function as well as reasoning. Path plots display standardized effects and p-values: (a) AF to imaging marker, (b) imaging marker to cognitive score, (ab) indirect effect (c') direct effect and (c) total effect. Significant associations are in blue; non-significant in light gray. If a relationship is significantly mediated, i.e., the indirect effect ab was significant and the direct effect c' was reduced or non-significant compared to the total effect c, the text for ab is highlighted in blue. The left path plots show results regarding attention/executive function, while the right path plots depict those of reasoning.

#### Figure S9 – Regression analysis: cognition ~ cortical macrostructural imaging markers

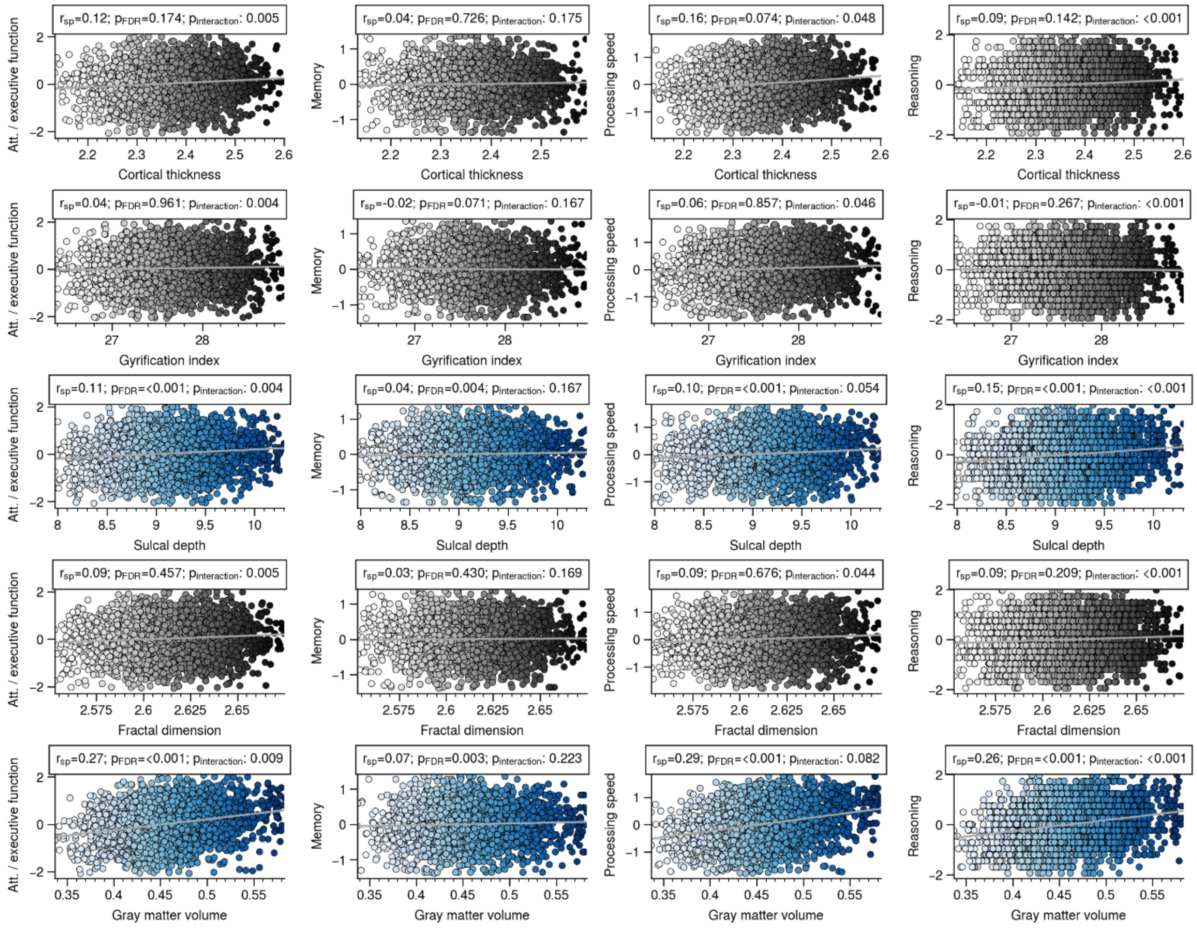


Linear regression analysis of the association between macrostructural cortical markers and cognitive outcomes. Analysis incorporated cognitive domain scores as the dependent variable, global imaging markers as the independent variable, and included age, sex, education and cardiovascular risk factors as covariates. The interaction effect of an atrial fibrillation diagnosis (global imaging marker * atrial fibrillation) was also tested. Significant associations between imaging and cognitive scores are denoted by blue dots. Non-significant associations are shown in gray. Abbreviations: $p_{FDR}$ – false discovery rate corrected p-value, adjusted for age, sex, education, and cardiovascular risk; $p_{interaction}$ - p-value for the interaction term (imaging marker * atrial fibrillation) with false discovery rate correction; $r_{sp}$ – Spearman correlation.

### Figure S10 – Regression analysis: cognition ~ cortical microstructural imaging markers

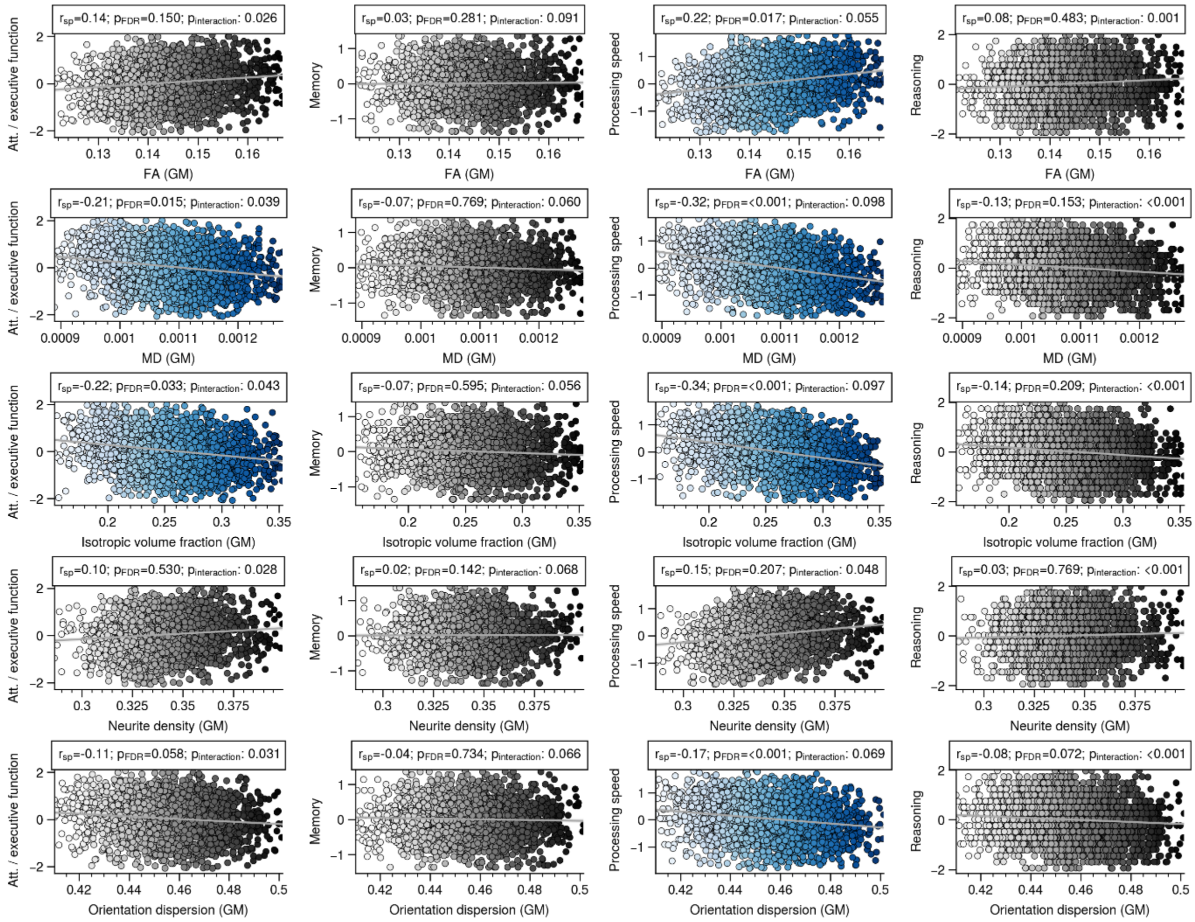


Linear regression analysis of the association between microstructural cortical markers and cognitive outcomes. Analysis incorporated cognitive domain scores as the dependent variable, global imaging markers as the independent variable, and included age, sex, education and cardiovascular risk factors as covariates. The interaction effect of an atrial fibrillation diagnosis (global imaging marker * atrial fibrillation) was also tested. Significant associations between imaging and cognitive scores are denoted by blue dots. Non-significant associations are shown in gray. Abbreviations: $p_{FDR}$ – false discovery rate corrected p-value, adjusted for age, sex, education, and cardiovascular risk; $p_{interaction}$ - p-value for the interaction term (imaging marker * atrial fibrillation) with false discovery rate correction; $r_{sp}$ – Spearman correlation.

#### Figure S11 – Regression analysis: cognition ~ white matter imaging markers

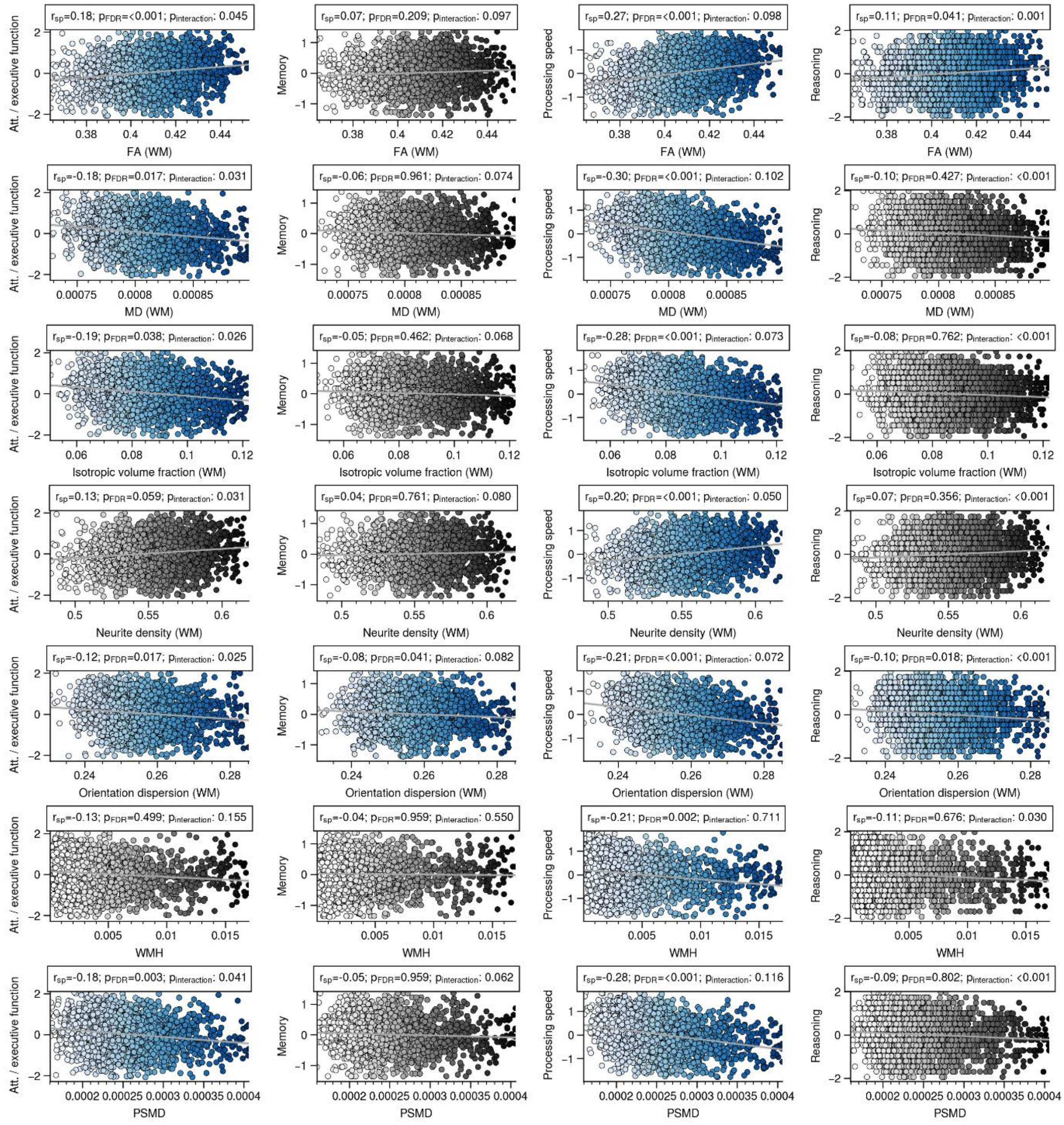


Linear regression analysis of the association between white matter markers and cognitive outcomes. Analysis incorporated cognitive domain scores as the dependent variable, global imaging markers as the independent variable, and included age, sex, education and cardiovascular risk factors as covariates. The interaction effect of an atrial fibrillation diagnosis (global imaging marker * atrial fibrillation) was also tested. Significant associations between imaging and cognitive scores are denoted by blue dots. Non-significant associations are shown in gray. Abbreviations: $p_{FDR}$ – false discovery rate corrected p-value, adjusted for age, sex, education, and cardiovascular risk; $p_{interaction}$ - p-value for the interaction term (imaging marker * atrial fibrillation) with false discovery rate correction; $r_{sp}$ – Spearman correlation.

### Figure S12 – Cortical effect maps comparison

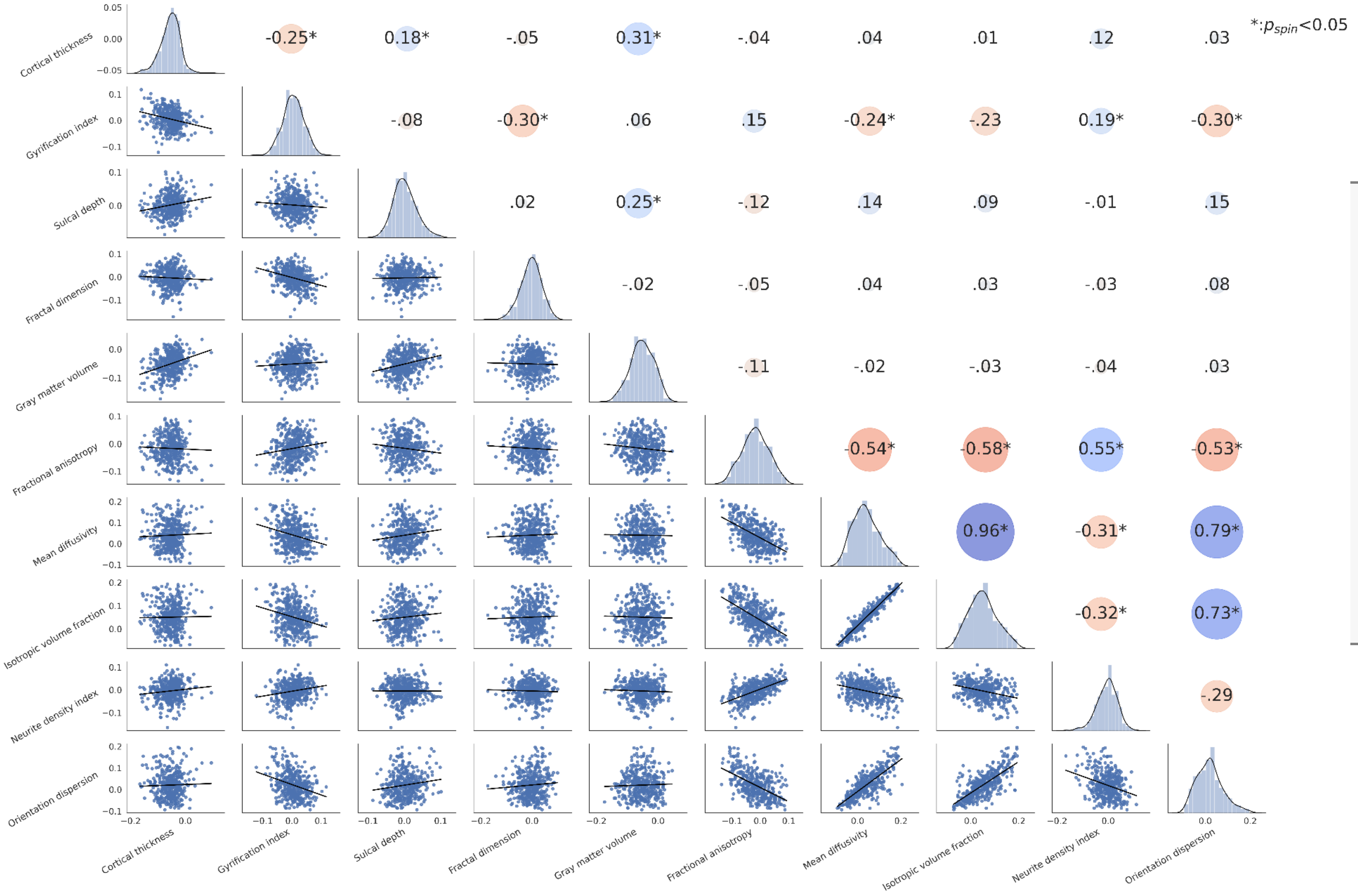


The correlation matrix displays spatial correlations of Schaefer400-parcellated Cohen's d maps resulting from the comparison of regional imaging markers between AF and controls. The lower triangle of the matrix displays scatter plots and the upper triangle the Pearson correlation coefficients of the corresponding correlations. The color and size of the dots on the upper triangle encode the effect directionality and effect size, respectively. The asterisks indicate statistical significance. To account for spatial smoothness, p-values were determined based on spin permutations ($p_{spin}$).[17] The matrix diagonal displays histograms.

### Figure S13 - Group comparison of tract-level fractional anisotropy (FA)

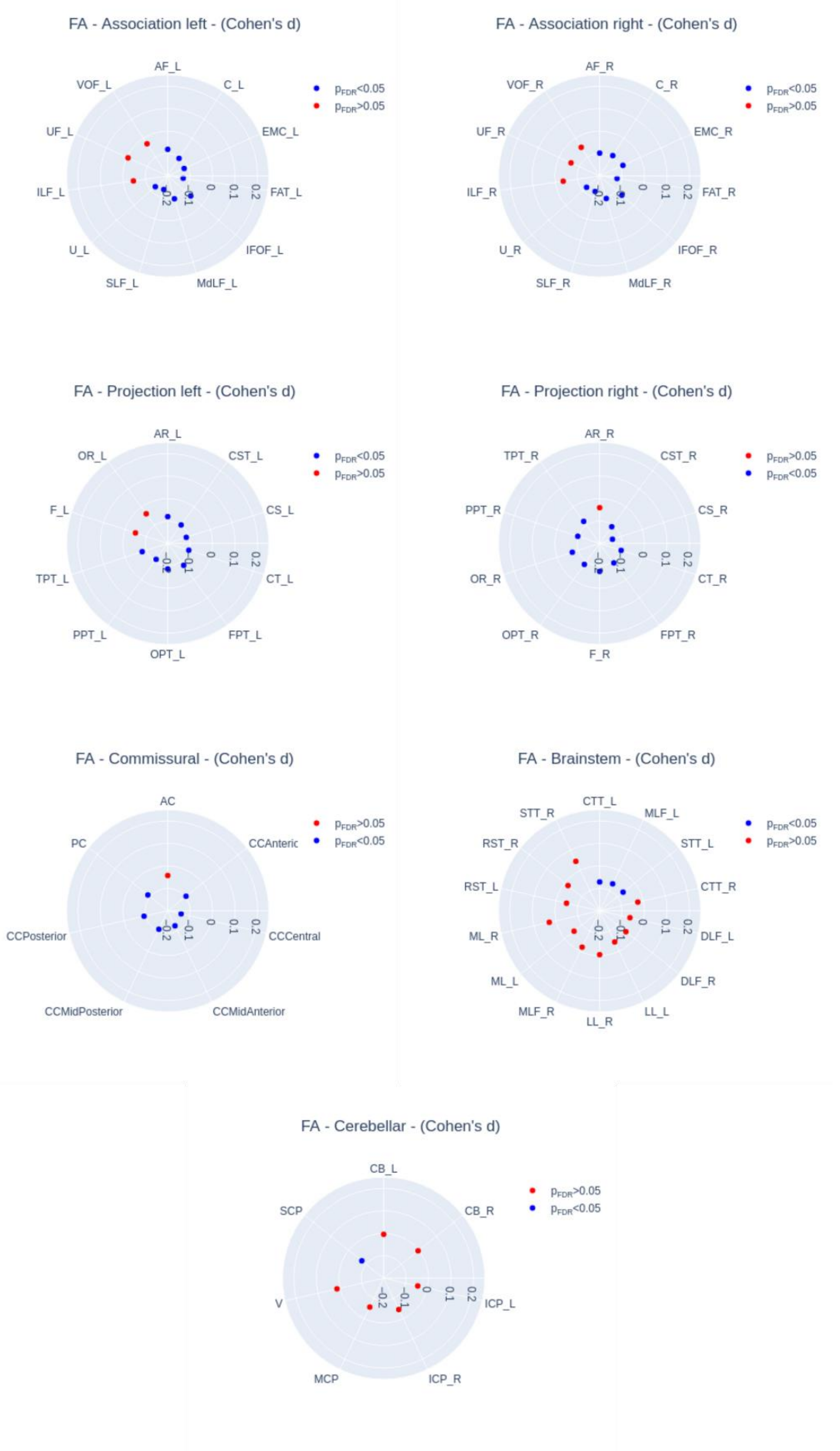


Radar plots represent the group differences for fractional anisotropy in each tract, with blue dots indicating significant differences and red dots marking non-significant differences.

### Figure S14 - Group comparison of tract-level mean diffusivity (MD)

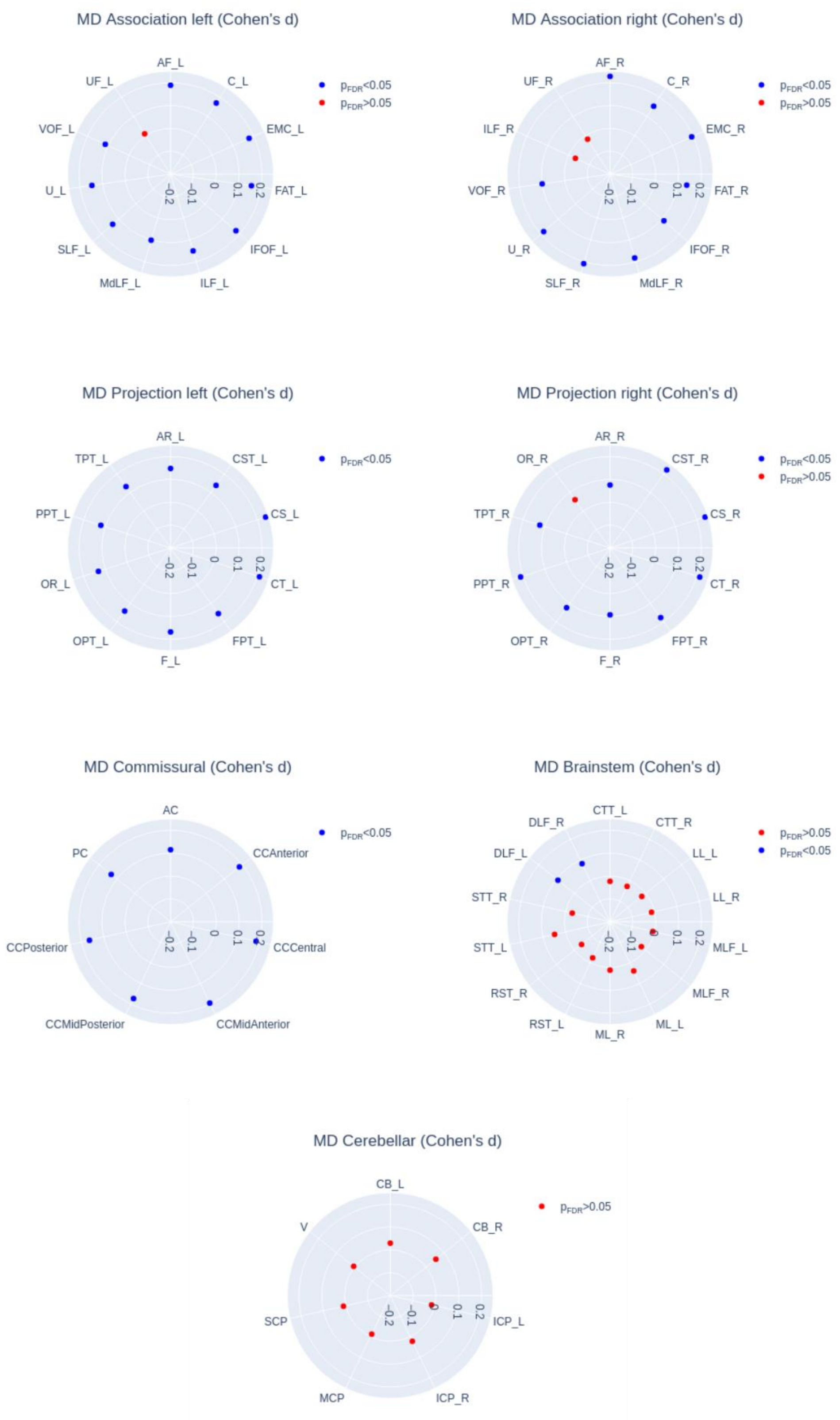


Radar plots represent the group differences for mean diffusivity in each tract, with blue dots indicating significant differences and red dots marking non-significant differences.

#### Figure S15 - Group comparison of tract-level neurite density index

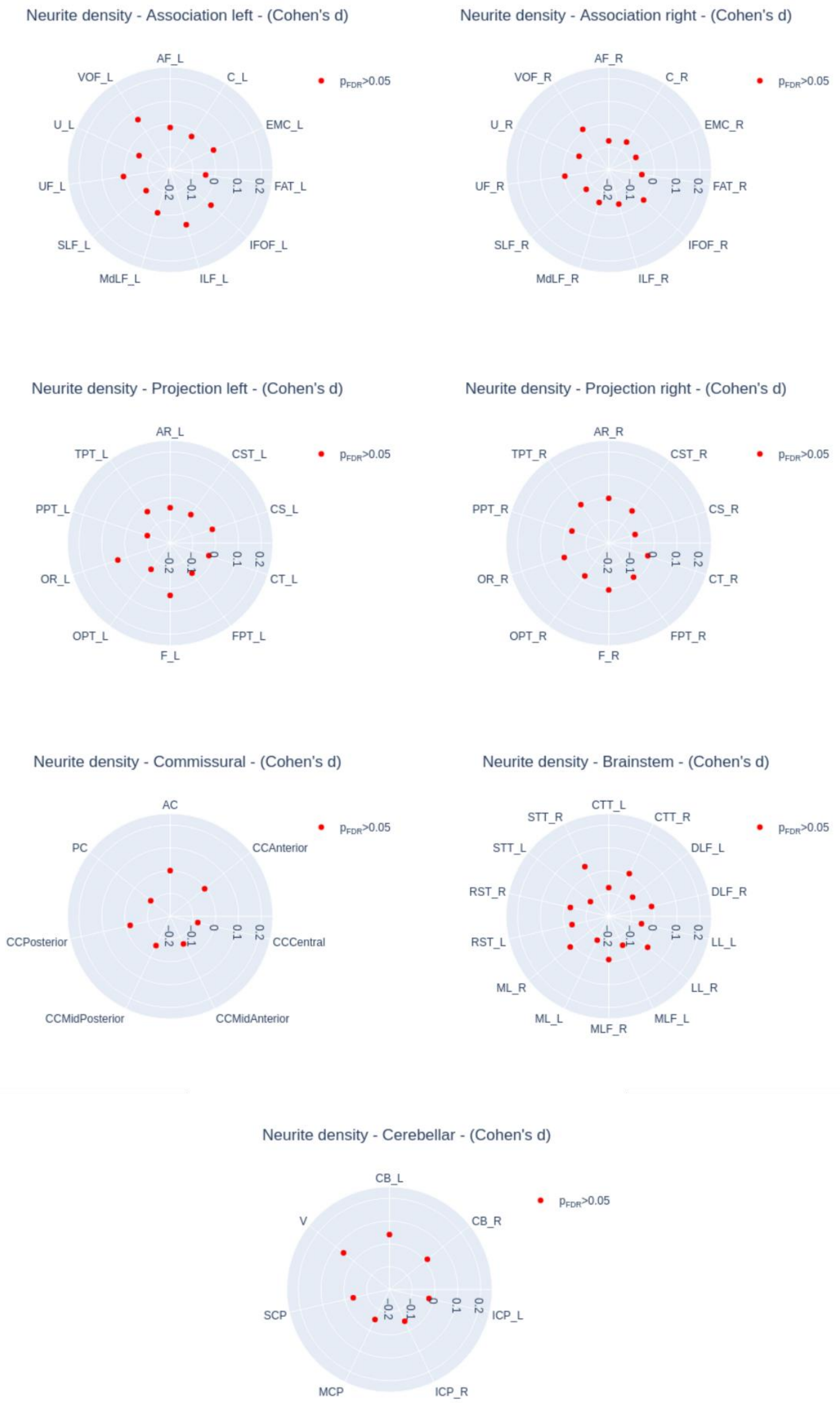


Radar plots represent the group differences of neurite density in each tract, with blue dots indicating significant differences and red dots marking non-significant differences.

#### Figure S16 - Group comparison of tract-level orientation dispersion

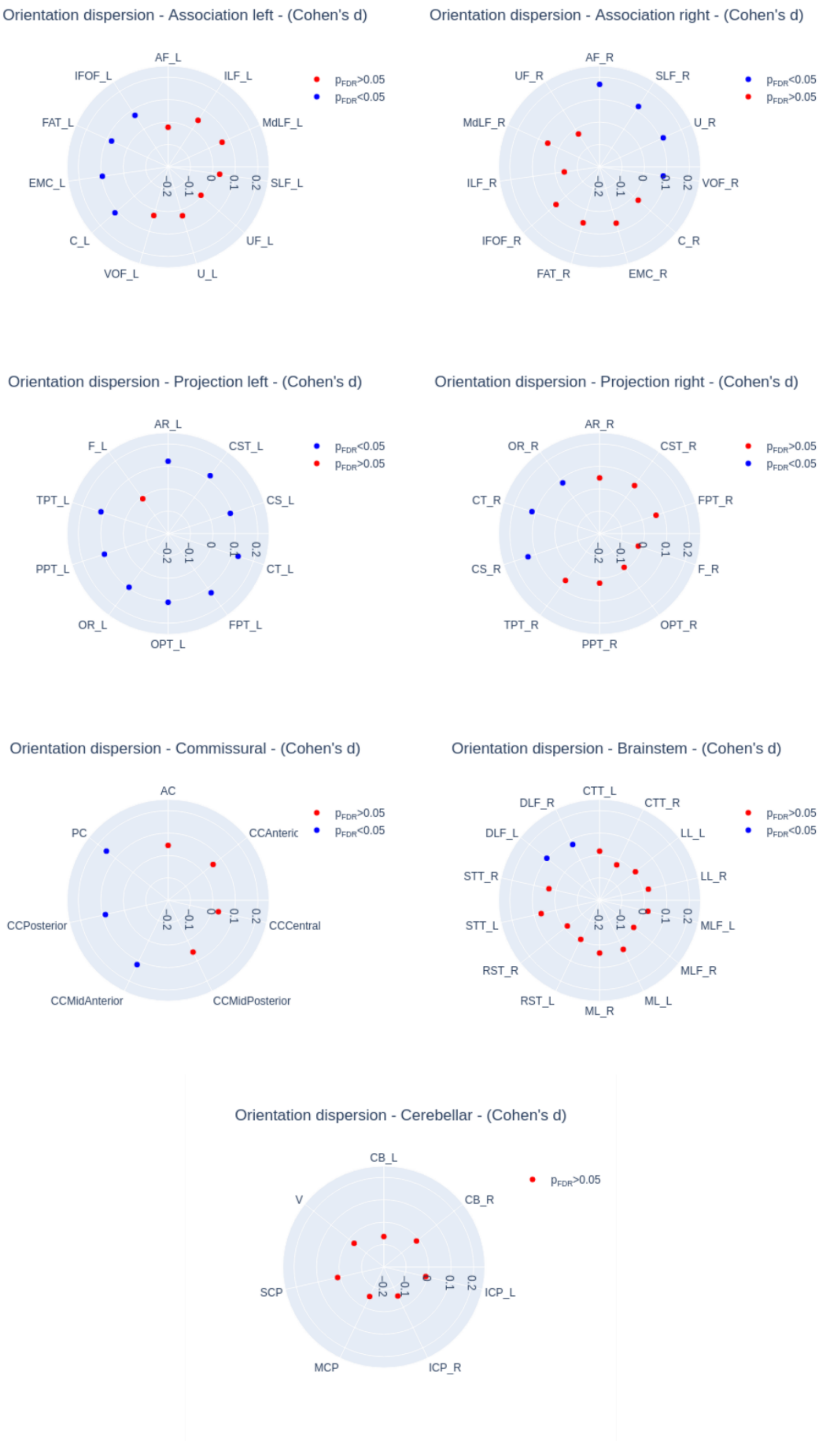


Radar plots represent the group differences for orientation dispersion in each tract, with blue dots indicating significant differences and red dots marking non-significant differences.
